# Sustaining effective COVID-19 control in Malaysia through large-scale vaccination

**DOI:** 10.1101/2021.07.05.21259999

**Authors:** Pavithra Jayasundara, Kalaiarasu M. Peariasamy, Kian Boon Law, Ku Nurhasni Ku Abd Rahim, Sit Wai Lee, Izzuna Mudla M. Ghazali, Milinda Abayawardana, Linh-Vi Le, Rukun K.S.Khalaf, Karina Razali, Xuan Le, Zhuo Lin Chong, Emma S McBryde, Michael T Meehan, Jamie M. Caldwell, Romain Ragonnet, James M Trauer

## Abstract

**Introduction:** As of 3^rd^ June 2021, Malaysia is experiencing a resurgence of COVID-19 cases. In response, the federal government has implemented various non-pharmaceutical interventions (NPIs) under a series of Movement Control Orders and, more recently, a vaccination campaign to regain epidemic control. In this study, we assessed the potential for the vaccination campaign to control the epidemic in Malaysia and four high-burden regions of interest, under various public health response scenarios.

**Methods:** A modified susceptible-exposed-infectious-recovered compartmental model was developed that included two sequential incubation and infectious periods, with stratification by clinical state. The model was further stratified by age and incorporated population mobility to capture NPIs and micro-distancing (behaviour changes not captured through population mobility). Emerging variants of concern (VoC) were included as an additional strain competing with the existing wild-type strain. Several scenarios that included different vaccination strategies (i.e. vaccines that reduce disease severity and/or prevent infection, vaccination coverage) and mobility restrictions were implemented.

**Results:** The national model and the regional models all fit well to notification data but underestimated ICU occupancy and deaths in recent weeks, which may be attributable to increased severity of VoC or saturation of case detection. However, the true case detection proportion showed wide credible intervals, highlighting incomplete understanding of the true epidemic size. The scenario projections suggested that under current vaccination rates complete relaxation of all NPIs would trigger a major epidemic. The results emphasise the importance of micro-distancing, maintaining mobility restrictions during vaccination roll-out and accelerating the pace of vaccination for future control. Malaysia is particularly susceptible to a major COVID-19 resurgence resulting from its limited population immunity due to the country’s historical success in maintaining control throughout much of 2020.

## Introduction

Malaysia is a multi-ethnic and populous country of 32.75 million people (Department of Statistics Malaysia, 2021) that has experienced three progressively worse waves of SARS-CoV-2 transmission to-date and has enforced a combination of non-pharmaceutical interventions (NPIs) and, more recently, vaccination roll-out to curtail transmission. The first imported cases of COVID-19 in Malaysia were detected in late January 2020 but case numbers remained low until a mass religious event held in Sri Petaling, Kuala Lumpur between 27 February and 1 March 2020 (Hashim *et al*., 2021; World Health Organization Representative Office for Malaysia Brunei Darussalam and Singapore, 2020). The case numbers were subsequently controlled through a nationwide movement control order (MCO) and a series of NPIs including physical distancing, mandated face coverings, limits on social gathering, inter-district travel bans and closure of schools and non-essential economic activities (World Health Organization Representative Office for Malaysia Brunei Darussalam and Singapore, 2021) (Fig. 1). Despite a less restrictive MCO (dubbed “MCO 2.0”) (Hashim *et al*., 2021; Teoh S, 2021), a second wave of transmission peaked in late January 2021 after previous MCO and NPIs were relaxed in early December 2020. Subsequently, Malaysia launched its National COVID-19 Immunization Programme with BNT162b2, followed by the addition of CoronaVac and ChAdOx1 (The Special Committee for Ensuring Access to COVID-19 Vaccine Supply (JKJAV), 2021). A third wave of COVID-19 is currently occurring (as of 3^rd^ June 2021) and may be attributable to easing of NPIs and social gatherings for the observation of religious festivals such as Eid. However, the third wave differs from the prior two waves in that three out of four currently circulating SARS-CoV-2 strains have been classified variants of concern (VoC: B.1.1.7, B.1.351 and, more recently, B.1.617) (World Health Organization, 2021; World Health Organization Representative Office for Malaysia Brunei Darussalam and Singapore, 2021). Restrictive NPIs have been reinstated along with a vaccination campaign to regain control. The MCO strongly impact society and the economy, with an estimated RM 2.4 billion lost per day (Shukry A, 2020). However, with little natural population immunity attributable to many months of well-controlled transmission, Malaysia is largely reliant on an effective vaccination strategy for the safe release of restrictions.

**Figure 1:**
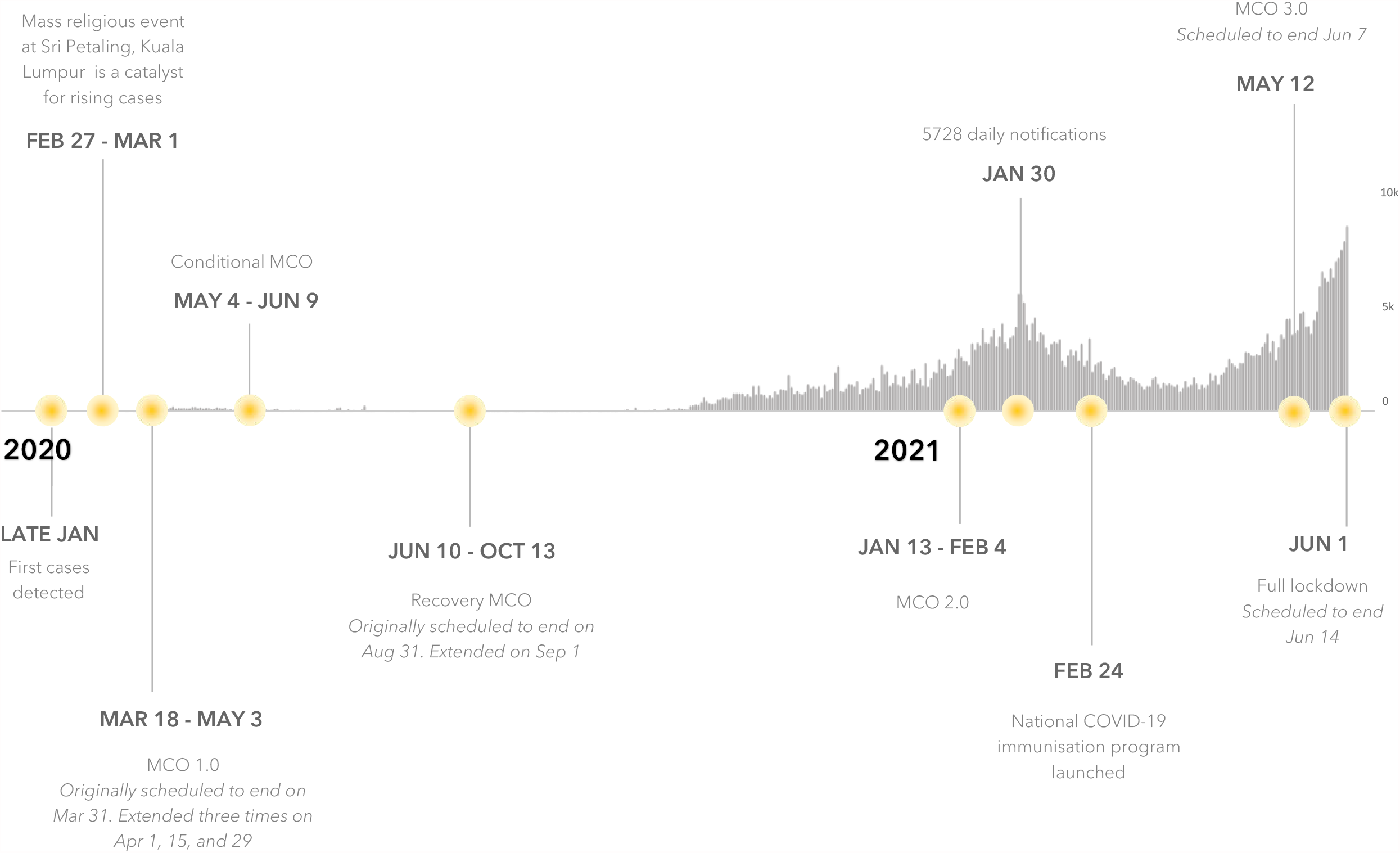
Summary of the COVID-19 epidemic in Malaysia and different movement control orders (MCO) introduced. MCO 1.0 restricted religious, social, educational and non-essential activities and inter-state travel. The conditional MCO (CMCO) partially allowed social activities and inter-state travel, and allowed non-essential economic activities. The CMCO was not implemented in all states. The recovery MCO (RMCO) partially allowed religious and educational activities and allowed inter-state travel. The MCO restrictions were reinstated in certain states for a second time in January 2021 and a third time in May 2021 (widely dubbed MCO 2.0 and MCO 3.0 respectively). During the full lockdown, all social and economic sectors are not allowed to operate except for essential services.

Malaysia currently has agreements for enough vaccine doses to cover 120% of the population (Bernama, 2021); however only a small proportion has been received as of May 2021, and with vaccination rates of 80,000 per day, the country faces challenges to meet its phase two targets (Ariff I., 2021). By the end of June 2021, 2.2 million doses of the BNT162b2 vaccine, 8.2 million doses of CoronaVac vaccine and 0.5 million doses of the ChAdOx1 vaccine are expected to have been administered in the National Covid-19 Immunisation Programme. In Q3 2021 Malaysia expects to receive a further 25 million doses of BNT162b2 vaccine, 4 million doses of CoronaVac and about 1 million doses of the ChAdOx1 vaccine. As of 26 May 2021, 6.7% of the population has received their first dose, whilst 3.8% have completed two doses (JKJAV Malaysia @ Twitter, 2021).

In this study, we model the impact of vaccination roll-out and relaxation of NPIs on the control of SARS-CoV-2 disease at the national level, and for four regions of interest (Kuala Lumpur, Selangor, Johor, Penang) selected due to the high number of cases and population density. Here, we model the COVID-19 epidemic in Malaysia to-date and consider the effectiveness of the vaccine program in the context of VoC and the social and economic imperatives to relax NPIs.

## Methods

### Model description

The application of a similar modelling framework is described elsewhere (Caldwell *et al*., 2021) and the full model description (including differential equations) is provided in the Supplementary File, with all code publicly available through https://github.com/monash-emu/AuTuMN/. The model consists of a modified susceptible (S)-exposed (E)-infectious (I)-recovered (R) compartment framework. The exposed category represents the incubation period and is sequentially divided into non-infectious exposed, and infectious exposed, while the infectious category is divided as ‘early active’ and ‘late active’ to capture incomplete case detection, isolation following detection and hospitalisation (Fig. 2). To account for age-specific differences in disease and differences in social-mixing, the base model is stratified by age into five-year bands from birth to ≥75 years, and age-structured heterogeneous mixing is implemented through a synthetic mixing matrix for Malaysia (Prem *et al*., 2017). We capture mobility through Google mobility data and incorporate “micro-distancing” which represents behaviours other than those that prevent people from directly coming into contact with one another, such as maintaining interpersonal physical distance and proper wearing of face masks.

**Figure 2:**
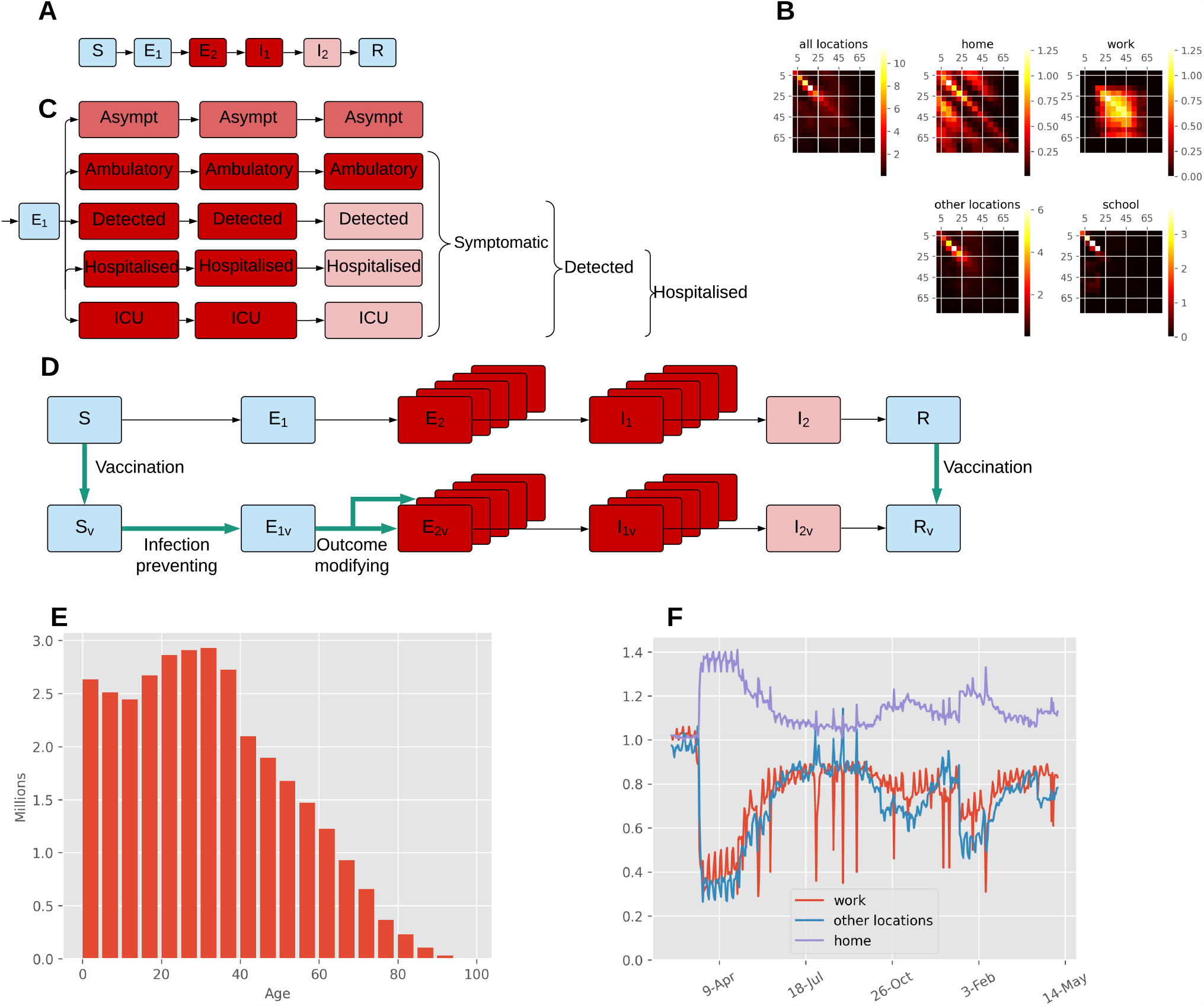
Illustration of key features of our age-structured COVID-19 model for Malaysia informed by population size, population mixing and mobility data. (A) Unstratified SEIR model structure, coloured by infectiousness of each state (blue = non-infectious; pink = moderately infectious; red = highly infectious). (B) Heterogeneous mixing matrices by age and location in the absence of non-pharmaceutical interventions (brighter colours indicate higher contact rates). (C) Stratification by clinical status (similar colour scheme to that in A). (D) Vaccine effects (turquoise bold arrows) for infection-preventing and severity-preventing vaccines. (E) Starting population age distribution. (F) Community quarantine driven mobility adjustments applied to the mixing matrices (before seven-day moving average smoothing).

### Model calibration

We calibrated the Malaysia national model using an adaptive Metropolis algorithm (Haario *et al*., 2001) to three targets: case notifications, intensive care unit (ICU) occupancy and infection-related deaths. Although the ICU occupancy and mortality data were too sparse for the regional models, aggregating the data to weekly national values allowed us to include these two targets (ICU occupancy and mortality) in the national model. Further details on the calibration procedure and the prior distributions of epidemiological calibration parameters are described in the Supplementary File and Table 1. We used uniform priors for highly uncertain quantities and truncated normal distributions for quantities informed by epidemiological evidence. As the prior distributions of the proportion of symptomatic individuals and the proportion of symptomatic individuals hospitalised are based on data from high income countries (HIC), we introduced adjuster parameters to account for any differences that could exist between these parameter values in HIC and low- and middle-income countries (LMIC) (which are multiplicative factors applied to the odds ratio corresponding to the adjustment being made, to ensure the resulting values remain proportions). The model was calibrated for the baseline scenario assuming the current mobility patterns as informed by Google mobility data remained in place from 3^rd^ June 2021 onwards. In the calibration process, we ran seven independent Adaptive Metropolis chains with ∼2000 iterations per chain and discarded the first 600 iterations as burn-in. Other than as described above, the model structure and calibration process were identical for the national model and the regional models.

**Table 1:**
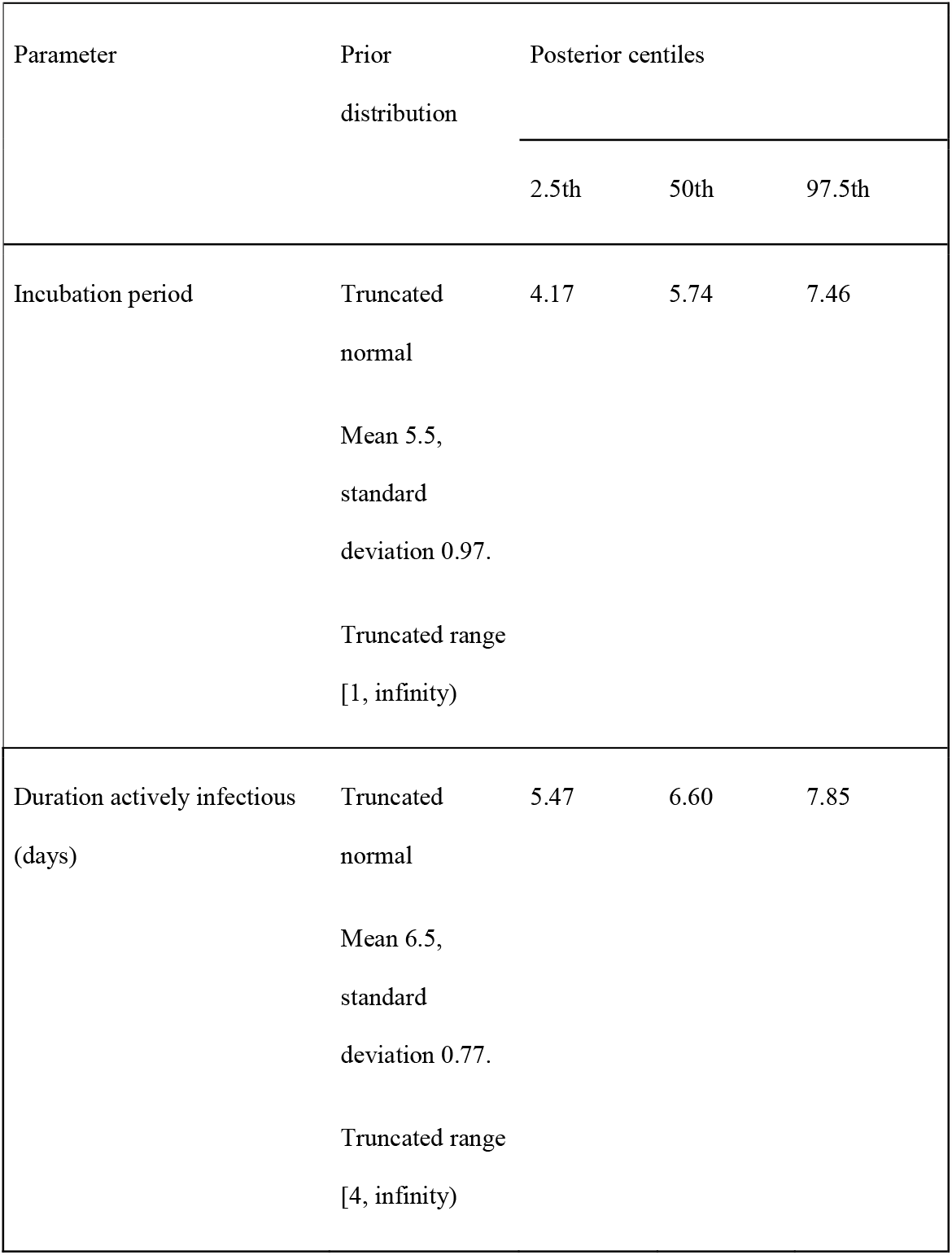
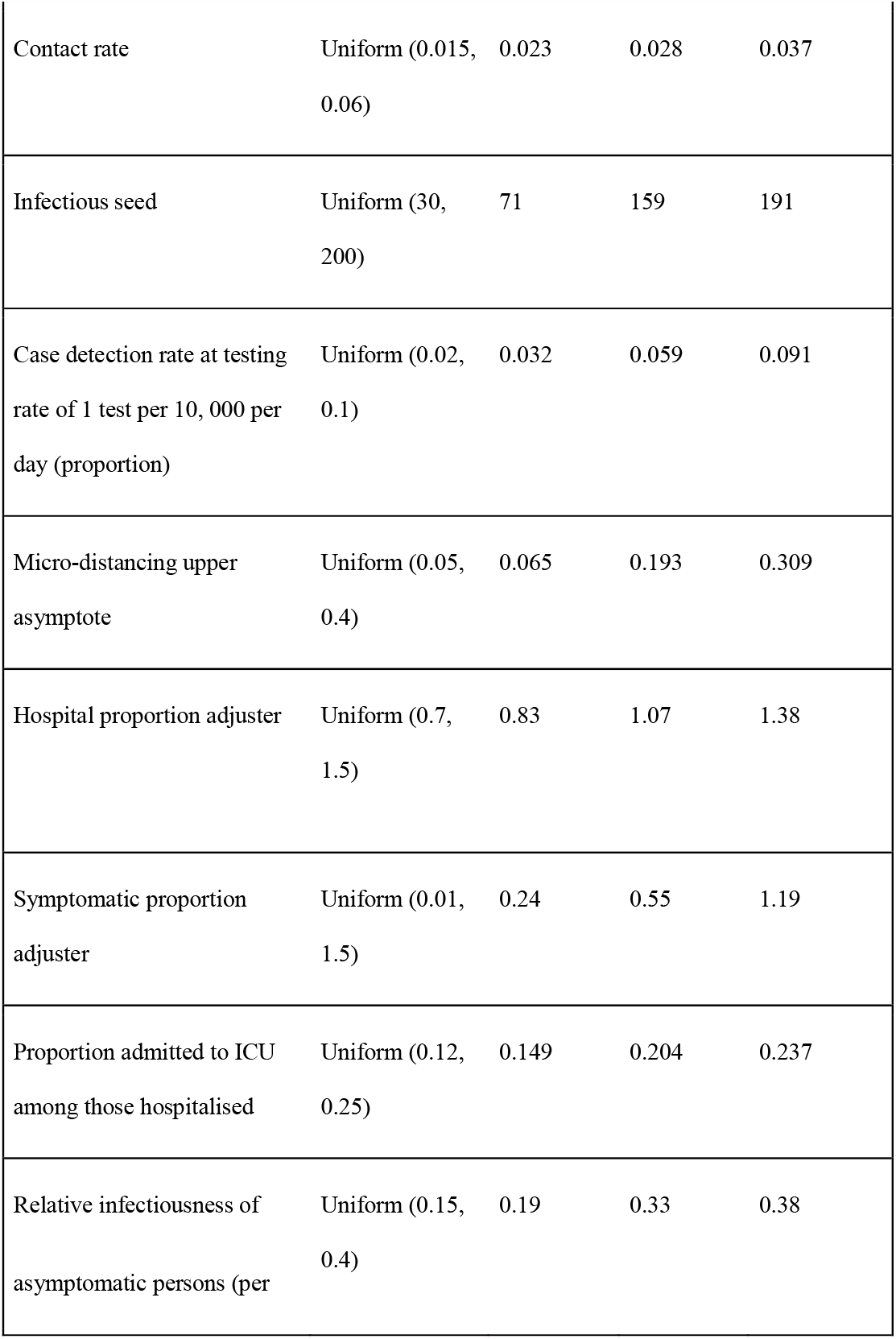
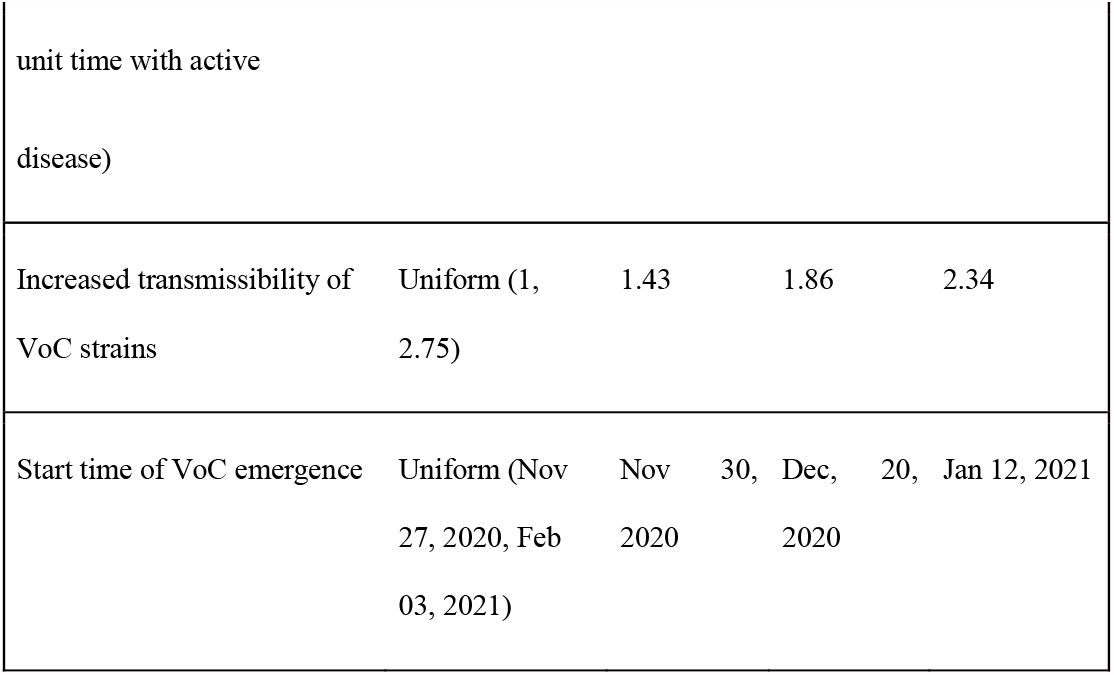
Prior and posterior distributions of all calibrated parameters

### Modelling variants of concern

To consider the effects of VoC on infection dynamics and vaccination programs, we explicitly simulated two competing strains to represent 1) the wild-type or ancestral virus, and 2) all VoC strains, where the VoC were assumed to be associated with increased transmissibility only. Susceptible individuals can be infected with either the wild-type or VoC strain and infectious individuals contribute to the force of infection with their respective infecting strain only. VoC strains are seeded into the model such that one additional person per day is infected with the VoC strain for a duration of ten days, with the time that this ten-day period commences varied during model calibration.

### Modelling vaccination

We stratified all model compartments as either vaccinated or unvaccinated and commenced simulations with a fully unvaccinated population. With vaccination roll-out, individuals in the susceptible and recovered compartments move from the unvaccinated to the vaccinated stratum at a constant rate. The daily rate of vaccination is calculated from the vaccine coverage achieved over a given period of time as 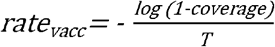, where coverage represents the final proportion of people vaccinated by the end of the roll-out period and *T* represents the roll-out period.

### Modelling vaccine effects

Vaccination is assumed to have two mechanisms of effect: 1) prevention of infection and 2) protection against progression to severe disease among those infected. A particular vaccine roll-out programme can be simulated to act through these two mechanisms simultaneously. We define the proportion of the effect that is attributed to preventing infection as 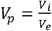, where *V*_*p*_ is varied through model calibration, *V*_*i*_ is the infection prevention efficacy and *V*_*e*_ is the overall efficacy against symptomatic disease (that would typically be the primary outcome of clinical trials). If severity prevention efficacy is denoted *V*_*s*,_ since *V*_*e*_*=V*_*i*_*+V*_*s*_*(1-V*_*i*_) it follows that 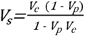 For the component of the vaccine effect attributed to infection prevention, the infection risk of vaccinated individuals is reduced by (*1-V*_*i*_). Severity-preventing vaccination reduces the infection fatality rate (IFR) and the probability that an infected individual experiences symptomatic disease (hence reducing the probability of hospitalisation). Thus, the vaccine efficacy parameter pertaining to disease severity prevention modifies the splitting proportions of infected individuals between the different clinical categories and the rate of COVID-19-related mortality.

### Modelling vaccination scenarios

We developed scenarios to consider the extent to which vaccination could permit the relaxation of NPIs, considering the extent to which vaccination protects against infection versus severe disease and the efficiency of the vaccination roll-out program. Scenarios considered were a return to the maximum mobility observed in the last 6 months (6 months since November, 2020) with roll-out of a 75% efficacious vaccine over 11 months to a final coverage of 80% (Scenario 1) and 50% (Scenario 2). These scenarios were chosen based on the vaccination targets that were deemed feasible by the study contributors from the Malaysia Ministry of Health. Because these vaccination projections were expected to lead to large epidemics that vaccination would be unable to substantially mitigate, we also combined these two vaccination scenarios with partial reductions in mobility, such that mobility values returned to a point part-way between current estimates and the maximum mobility observed in the last 6 months (creating four additional scenarios).

### Sensitivity analysis

We quantified the importance of various vaccination program-related parameters on the epidemic size over coming months by varying these parameters along with the calibration parameters in our scenario projections, under the three levels of mobility restrictions introduced above (full return to maximum recent mobility, 25% return and 50% return). We used partial rank correlation coefficients to determine the parameters that are most influential to the reduction in incidence from the baseline values two months after the start of interventions (referred hereafter as ‘relative incidence’). Here, the parameter values were randomly sampled for vaccination coverage ∼ uniform (0, 1), overall efficacy ∼ uniform (0, 1), *V*_*p*_ ∼ beta (13.58, 5.82) (the parameters of the beta distribution were chosen to be associated with an expectation of 70% and 95% of the distribution in [50%, 90%]) and other parameters are as described in Table 1.

## Results

### Parameter estimates

The posterior estimates of model calibration parameters are presented in Table 1 and the prior and posterior distributions are shown in Supplementary File, Fig. 4. The posterior distribution of the incubation period and infectious period were consistent with our prior beliefs informed from published literature. The posterior distribution associated with micro-distancing suggests it may have had a substantial effect in controlling the epidemic and this observation was consistent across all regions, with the impact from micro-distancing particularly strong in Johor (Supplementary File, Fig.5) where a particularly dramatic reversal in epidemic trajectory had been achieved by January, 2021. The increased transmissibility of VoC strains was substantially informed by the fitting process and was estimated at 1.86 (95% range: 1.43 - 2.34), implying that VoC are 86% more transmissible than the wild type strain which is consistent with the published estimates (Davies *et al*., 2021). The posterior density of VoC transmissibility at a value of one (implying equivalent transmissibility of all strains) was zero, suggesting that the recent surge in cases may be attributed to VoC circulation. The estimated start time of VoC emergence in Malaysia was early January 2021. The adjuster parameter that was used to account for any differences between HIC and LMIC for proportion symptomatic was < 1, suggesting this quantity to be likely lower in Malaysia than in the countries in which the baseline parameter was estimated (predominantly HIC, Supplementary File, Fig. 4). For the proportion of infected individuals hospitalised, there was no strong evidence to suggest that this was different in Malaysia compared to HIC. At around September 2020 the proportion of symptomatic cases detected is estimated at around 8% to 24% and this proportion gradually increased and currently (at 3^rd^ June, 2021) is estimated at 49 to 88% (Supplementary File, Fig. 7), although this was associated with considerable uncertainty.

**Figure 3:**
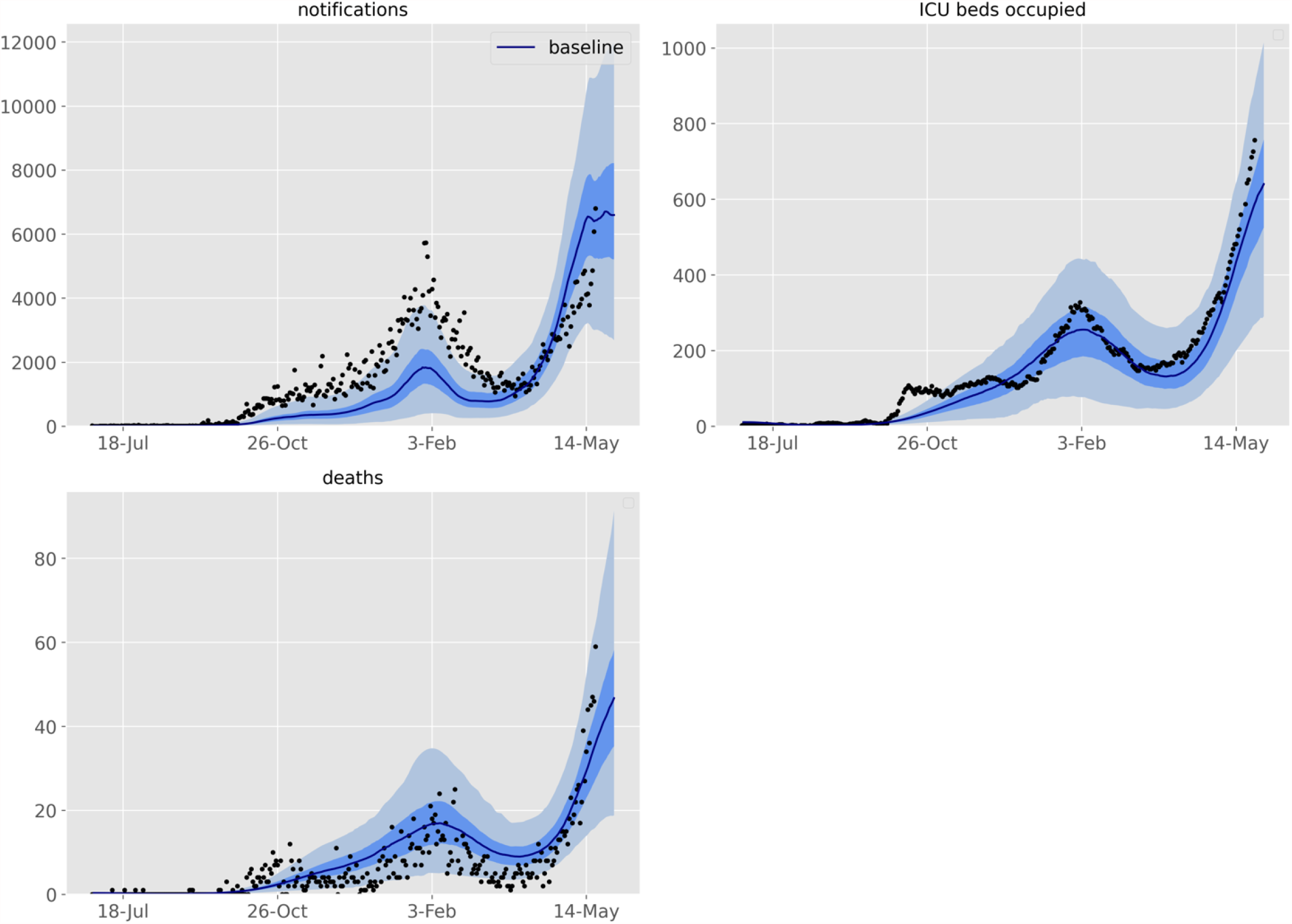
The Malaysia national model calibration fits for notifications, ICU occupancy and deaths. Line, 50th centile credible interval; dark shading, 25th to 75th centile credible interval; light shading 2.5th to 97.5th centile credible interval. Black circles; reported data.

**Figure 4:**
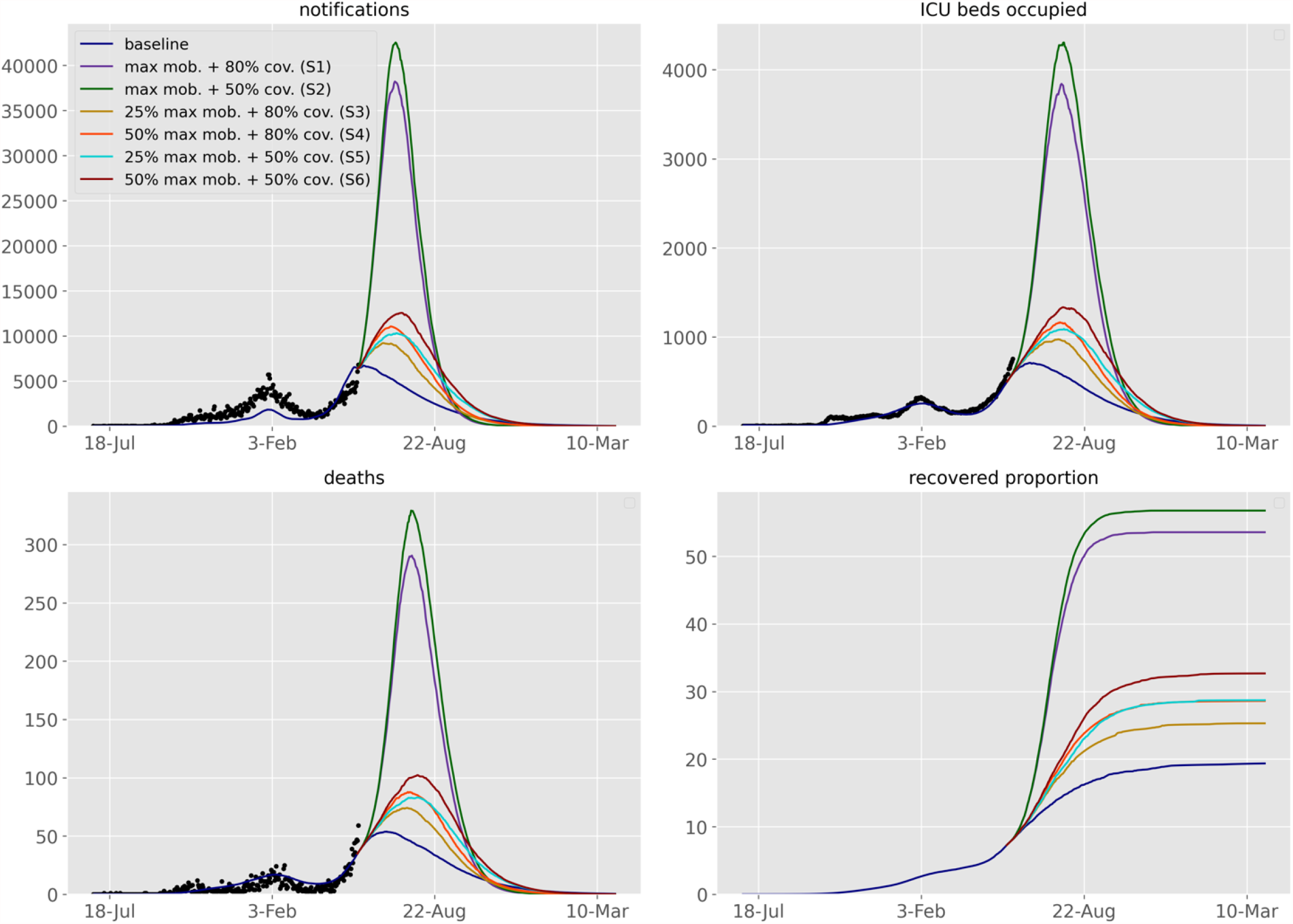
Future projections of the COVID-19 epidemic in Malaysia under various response scenarios and baseline. Upper-left, daily number of notifications; upper-right, ICU beds occupied; lower-left, daily number of COVID-19-related deaths; lower-right, proportion of recovered population. For better visualisation, the median fits and projections are shown without uncertainty bounds. The black scatters are the data points. The scenarios considered are, return to the maximum mobility with 80% (S1) and 50% (S2) vaccine coverage, 25% (S3) and 50% (S4) return to the maximum mobility with 80% vaccine coverage, 25% (S4) and 50% (S5) return to the maximum mobility with 50% vaccine coverage and 50% return to the maximum mobility with 50% vaccine coverage (S6).

### The epidemiological fit

For the national model and all four regional models we achieved a good fit to epidemiological indicators (especially case notifications) through a combination of time-varying processes that include changes to case detection rates, population mobility, physical distancing, micro-distancing and inclusion of VoC (national model fits shown in Fig. 3 and regional model fits in Supplementary File, Fig. 8). However, the national model underestimated the number of deaths and ICU occupancy over recent weeks. Because this change was so recent and further data continues to emerge, we did not further manipulate the model structure to capture these differences.

### Model validation

We validated our model using the data on cumulative notifications for the national model by different age groups and the model estimates align well with the observed data (Supplementary File, Fig. 9).

### Effect of vaccination strategies

To inform evidence-based relaxation of NPIs, we considered the impact of several vaccination strategies in controlling the epidemic in Malaysia (Fig. 4). If the current significant community control restrictions are maintained in the absence of vaccination, new COVID-19 notifications are projected to continue to increase, reaching a peak of ∼7250 cases around the beginning of June, 2021 (Fig. 4). Under this baseline, at the peak of the epidemic ∼60 daily deaths would occur and demand for around 980 ICU beds is predicted. With the relaxation of NPIs to the maximum mobility observed in the last 6 months and with vaccination coverage of 50% and 80% achieved over 11 months, a major epidemic is anticipated, with case numbers peaking around 8-fold and 7-fold higher than the previous peak in January 2021, respectively.

If a 25% or 50% return to the maximum mobility is maintained, both vaccination strategies of 50% and 80% coverage are substantially more successful in mitigating the projected epidemic, although the epidemic waves are still predicted to be large and markedly exceed the previous epidemic in January 2021. All these scenarios would result in a peak between June 20^th^ and July 5^th^, 2021. Strategies that reduce mobility and increase vaccine coverage are more effective in controlling the epidemic and among the tested scenarios, the scenario with 25% return to the maximum mobility and 80% vaccine coverage (Scenario 3) was most effective in controlling the epidemic. Under Scenario 3, peak case numbers are predicted to be 1.8-fold higher than the January, 2021 wave. During this peak, Scenario 3 may result in ∼75 median daily deaths (95% range: ∼20 - 200) and demand for median ICU occupancy of ∼1000 (95% range: ∼250 - 2500), which is likely to exceed the ICU capacity allocated for COVID-19 in Malaysia (1114 beds).

Overall these results indicate that even with successful vaccination strategies, in order to reduce the COVID-19 case numbers in Malaysia, sustained mobility restrictions are likely to be required, with this being a consistent finding in both national and regional models other than Johor (Supplementary File, Fig. 10-13). In Johor, except under the maximum mobility scenarios, better epidemic control may be achievable through vaccination. This finding may be attributable to the relatively modest increased transmissibility of VoC estimated for Johor and the substantial impact of micro-distancing (Supplementary File, Fig. 6). However, as VoC replace the wild-type virus across the country, VoC may have increasingly consistent effects across regions. The median proportion of recovered individuals in the national model under scenarios 1, 2, 3, 4, 5 and 6 were estimated at approximately 0.52, 0.55, 0.25, 0.29, 0.29 and 0.32, respectively (Fig. 4).

**Figure 5:**
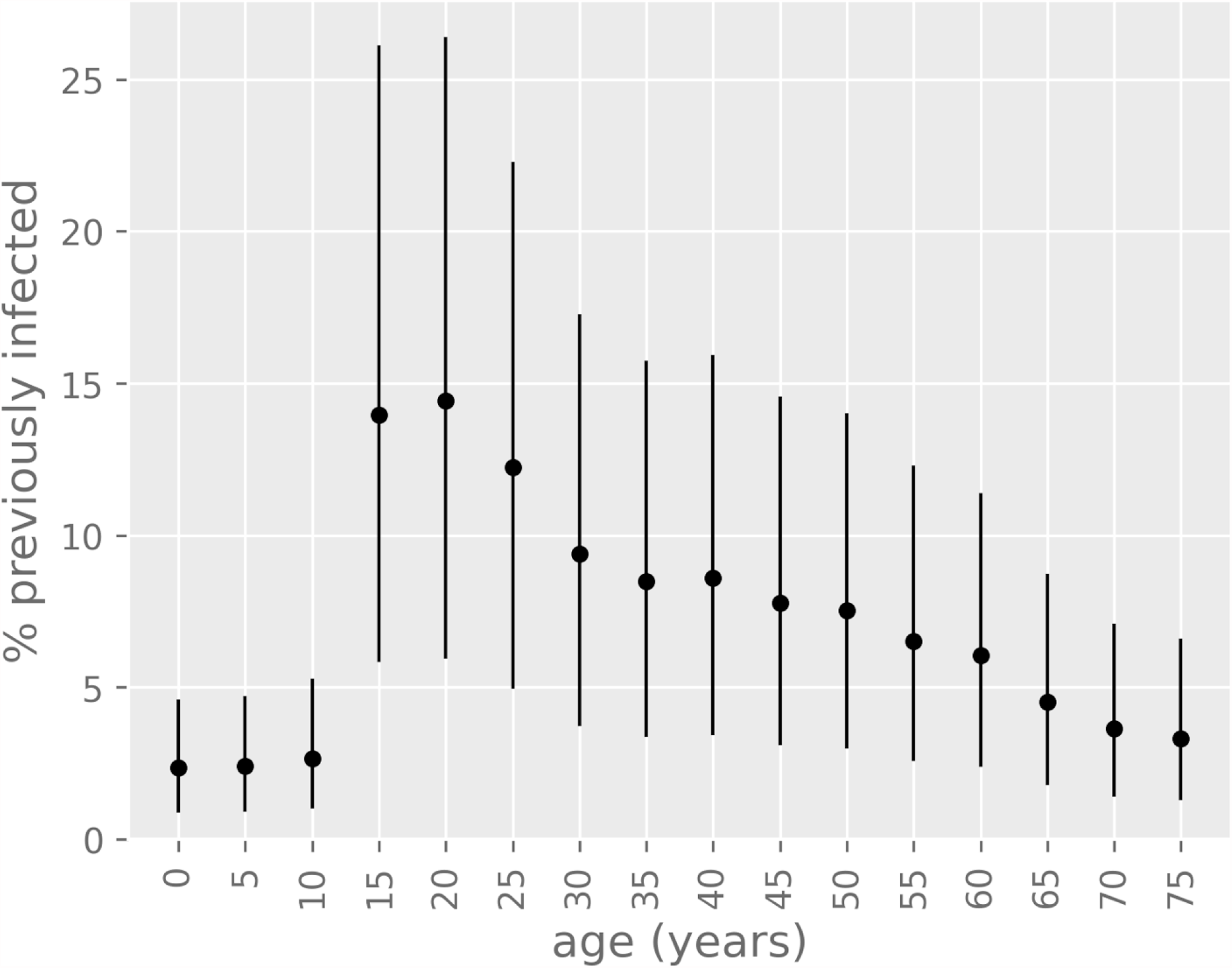
Percentage of previously infected individuals (modelled) by age distribution. Dots represent median estimates and whiskers represent the 95% credible interval. Results are shown for the baseline scenario as of 3^rd^ June, 2021.

**Figure 6:**
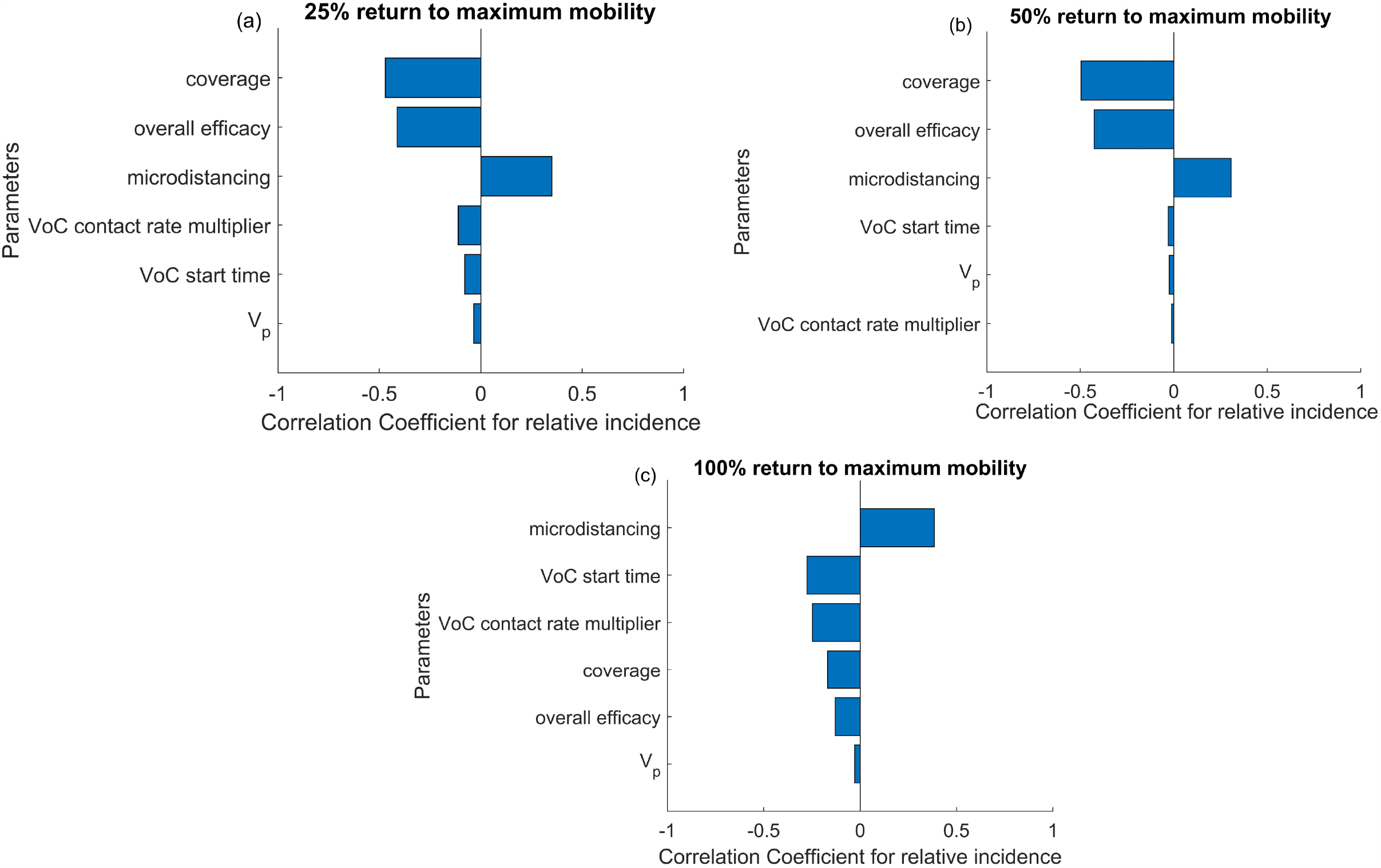
Tornado plots of partial rank correlation coefficients, indicating the importance of each parameter’s uncertainty in contributing to the change in incidence after two months. Sensitivity analysis was conducted under the three background mobility levels of (a) 25%, (b) 50% (b) and (c) 100% return to the maximum mobility observed in the past 6 months.

### The impact of different age groups

All models predicted that individuals aged between 15-25 years contributed to the highest proportion of infections with the seroprevalence of these age groups ranging ∼5% – 27% as of 3^rd^ June, 2021 (Fig. 5). However, there was considerable uncertainty around the absolute extent of population immunity.

### Sensitivity analysis

Vaccine coverage and overall efficacy were the most important contributors to the variability in the relative incidence under the mobility levels of 25% and 50% return to the maximum mobility observed in the past 6 months (Fig. 6). Micro-distancing was found to be an important predictor under all three background mobility assumptions. Under the maximum-mobility scenario, we found that vaccination had insufficient opportunity to control the large and rapid epidemic wave, such that maintaining micro-distancing was more influential.

## Discussion

We were able to capture the profile of Malaysia’s main COVID-19 epidemic wave using a model that incorporates age-specific infection, various clinical outcomes and Google mobility data. However, the true size of the epidemic was relatively poorly constrained through the calibration process due to uncertainty concerning the proportion of COVID-19 episodes identified. This emphasises the importance of other objective measurements of the epidemic size, such as sero-surveys, which in Malaysia are only available from prior to the major epidemic waves we simulated (personal correspondence, Zhuo Lin Chong). Our scenario projections indicated that complete release of all NPIs would result in a major resurgence of cases that would be negligibly mitigated by vaccination due to the relatively slow vaccination roll-out and concurrent circulation of VoC. Therefore, even with accelerated vaccination roll-out, the epidemic is unlikely to be controlled unless substantial mobility restrictions are continued. This is further emphasised by the finding that even if restrictions are partially maintained, vaccination would be unable to prevent an epidemic far beyond those previously experienced in Malaysia. Despite notably different epidemic profiles in the sub-regions modelled, results were generally consistent for each of the four Malaysian Provinces simulated, except in Johor. In reality, full VoC strain replacement may not yet have occurred in Johor, which will become clearer in the coming weeks to months. Based on these findings, when implementing public health guidelines, the model results suggest the importance of maintaining micro-distancing and mobility restrictions while carrying out the vaccination program in order to control the COVID-19 pandemic and prevent increases in COVID-19 related deaths and hospitalisations in the next few months.

The cause of the recent surge in cases observed in Malaysia is incompletely understood partially due to limited genomic testing that is often targeted to local public health priorities. However, from the limited testing available, patterns from several other countries indicating replacement of the wild-type strain with VoC (Williams *et al*., 2021) and discussions with in-country staff, VoC are currently considered the most likely explanation. Three VoC have been identified in Malaysia since mid-March (B.1.1.7, B.1.351 and B.1.617); timing that would be consistent with VoC being responsible for the recent resurgence in cases. Nevertheless, because of the significant uncertainty around the current epidemiology and the recency of the resurgence in case numbers, we sought to avoid over-fitting to limited data. We therefore implemented a model of VoC emergence in which VoC were distinguished by their increased transmissibility only. Our projections underestimated ICU occupancy and deaths (Fig. 3) in recent weeks compared to case notifications, whereas the reverse profile was observed in the earlier phases of the epidemic. This may be attributable to increased severity of VoC (Freitas *et al*., 2021; Iacobucci, 2021), or to saturation of case detection (Batumalai K, 2021; Tan *et al*., 2021), which are not captured by the current model configuration. Currently, in Malaysia 25% of detected cases are unlinked to known clusters (Ministry of Health Malaysia, 2021). Therefore, VoC are likely to hinder the relaxation of NPIs, as the model simulations predict a large epidemic if mobility restrictions are relaxed. This emphasises the need for sustained mobility restrictions and increased control measures to limit the spread of COVID-19 cases in Malaysia while the vaccination program is delivered. Similar findings of the need for maintaining control measures while carrying out vaccination programs in the presence of VoC have been reported for the United States (Borchering RK, 2021) and Italy (Giordano *et al*., 2021).

In the model, we considered vaccination scenarios that achieved 50% and 80% coverage over 11 months, targets that were deemed feasible by study contributors from the Malaysia Ministry of Health. However, under these strategies our projections suggest a need to maintain substantial NPIs for much of the roll-out period to control the epidemic. Therefore, in order to return to normal societal and economic functioning more quickly, it will be necessary to substantially accelerate the vaccination program from what is currently deemed feasible. This highlights the potential benefits of accelerated vaccination, which would require a focus on all aspects of the program, including procurement, logistics, public confidence and engagement of marginalised groups. Without such acceleration, the country likely faces the choice between a need for substantial NPIs for several months to come or a major outbreak likely to overwhelm health services. Several steps are currently being taken to achieve accelerated roll-out and the vaccination rate is expected to increase substantially from July, with confirmed vaccine delivery, more than 600 vaccination centres and the anticipated official approval of additional vaccines.

The negligible modelled population recovered from infection (<0.2%) (Fig. 4) aligns with a national seroprevalence study conducted from 7th August to 11th October, 2020 (personal correspondence, Zhuo Lin Chong) which reported a seroprevalence of 0.3% in those aged under 18 and 0.7% among those aged 18 and above. However, it remains difficult to determine the true size of the epidemic in the absence of recent serosurveillance data for validation, which has a significant impact on the projected future epidemic size through the simulated size of the recovered population.

The model is subject to several limitations especially when predicting over a long time-frame (Berger *et al*., 2021), only some of which are accounted for through the uncertainty ranges allowed around calibration parameter values. In particular, it is impossible to predict the extent to which NPIs will be maintained, which is dependent on societal and economic considerations in addition to health concerns. Our assumption that case detection rate scales with testing numbers does not capture the likely decrease in the efficiency of case ascertainment and contact tracing that likely occurs as incidence increases. The inclusion of VoC remains an assumption as described above. Our stratification by vaccination status does not include explicit states to differentiate between those who have received one or two doses of a two-dose regimen. Therefore, we could not consider the policy alternatives of maximising coverage with a single dose versus ensuring more people complete the full two-dose course. We did not simulate past vaccination roll-out because we assumed that vaccination has had a negligible effect to-date, given that only 3.8% of the targeted population have received both vaccine doses while 6.7% have received a single dose as of 24^th^ May, 2021 (JKJAV Malaysia @ Twitter, 2021). Although the second dose of BioNTech-Pfizer appears to result in ∼15% increase in effectiveness (Public Health England, 2021), insufficient data are currently available to compare the effectiveness between the doses of other vaccine options used in Malaysia. We also do not consider the effects of waning immunity (both vaccine-induced and infection-induced), although this may be important to consider in future implementations as several studies suggest waning antibody levels in individuals recovered from COVID-19 (Ward *et al*., 2020) and reports of repeated infection (Law S.K. *et al*., 2020). We also do not consider antigenic drift (Williams and Burgers, 2021) and immune escape (Kupferschmidt, 2021) which would be difficult to predict with existing data.

## Conclusion

Despite - and because of - good control throughout much of 2020, Malaysia faces the potential for a major epidemic of COVID-19 over the coming months. Although the true epidemic size to-date is incompletely understood, it is likely that the large majority of Malaysia’s population remain fully susceptible to infection. Further, strain replacement with VoC of increased transmissibility are the most likely explanation for the current resurgence in case numbers that has necessitated the recent return to lockdown. Therefore, rapid roll-out of vaccination is particularly critical to controlling the country’s epidemic, with continuing NPIs likely to be needed throughout much of the roll-out period.

## Data Availability

The data analysed during the current study and model code are publicly available.

https://github.com/monash-emu/AuTuMN/

## Acknowledgements

We would like to thank the Director General of Health Malaysia for his permission to publish this article. We also gratefully acknowledge the support of the Malaysian Ministry of Health. We also thank Amazon Web Services who provided the high-performance computing resources.

## Abbreviations

MCO: movement control order
ICU: intensive care unit
NPI: non-pharmaceutical interventions
VoC: variants of concern

