## Supplementary File for "Sustaining effective COVID-19 control in Malaysia through large-scale vaccination"

### Supplemental material supporting "Sustaining effective COVID-19 control in Malaysia through large-scale vaccination"

### 1 Base model construction

#### 1.1 Platform for infectious disease dynamics simulation

We developed a deterministic compartmental model of COVID-19 transmission using the AuTuMN platform, publicly available at https://github.com/monash-emu/AuTuMN/. Our repository allows for the rapid and robust creation and stratification of models of infectious disease epidemiology and includes pluggable modules to simulate heterogeneous population mixing, demographic processes, multiple circulating pathogen strains, repeated stratification and other dynamics relevant to infectious disease transmission. The platform was created to simulate TB dynamics, being an infectious disease whose epidemiology differs markedly by setting, such that considerable flexibility is desirable [1]. We have progressively developed the structures of our platform over recent years, and further adapted it to be sufficiently flexible to permit simulation of other infectious diseases for the purpose of this project.

#### 1.2 Base COVID-19 model

Using the base framework of an SEIR model (susceptible, exposed, infectious, removed), we split the exposed and infectious compartments into two sequential compartments each (SEEIIR). The two sequential exposed compartments represent the non-infectious and infectious phases of the incubation period, with the latter representing the "presymptomatic" phase such that infectiousness occurs during three of the six sequential phases. For this reason, "active" is a more accurate term for the two sequential "I" compartments and is preferred henceforward. The two infectious compartments represent early and late phases of active disease, during which symptoms occur if the disease episode is symptomatic, and allow explicit representation of notification, case isolation, hospitalisation and admission to ICU. The "active" compartment also includes some persons who remain asymptomatic throughout their disease episode, such that these compartments do not map directly to either persons who are infectious or those who are symptomatic (Figure 1).

The latently infected and infectious presymptomatic periods together comprise the incubation period, with the incubation period and the proportion of this period for which patients are infectious defined by input parameters described below. In general, two sequential compartments can be used to form a gamma-distributed profile of transition to infectiousness following exposure if the progression rates for these two compartments are equal, although in implementing this model the relative sojourn times in the two sequential compartments usually differed. Nevertheless, the profiles implemented are broadly consistent with the empirically observed log-normal distribution of individual incubation periods [2].

The transition from early active to late active represents the point at which patients are detected (for those persons for whom detection does eventually occur) and isolation then occurs from this point forward (i.e. applies during the late disease phase only, see Section 2). This transition point is also intended to represent the point of admission to hospital or transition from hospital ward to intensive care for patients for whom this occurs (see Section 1.4).

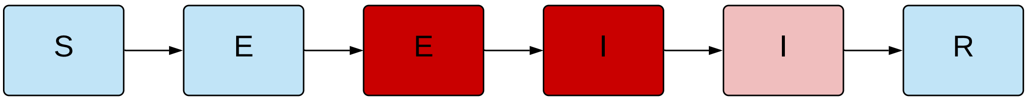

**Figure 1 – Unstratified compartmental model structure.** S = susceptible, E = exposed, I = active, R = recovered/removed. Depth of pink/red shading indicates the infectiousness of the compartment.

##### *1.3 Age stratification*

All compartments of this base compartmental structure were stratified by age into five-year bands from 0-4 years of age through to 70-74 years of age, with the final age group being those aged 75 years and older. Heterogeneous baseline contact patterns by age were incorporated using age-specific contact rates estimated by Prem et al. 2017 [3], who combined survey response data with information on national demographic characteristics to produce age-structured mixing matrices with these age groupings. These are then modified by non-pharmaceutical interventions as described in Section 3. Our modelled age groups were chosen to match these mixing matrices. The automatic demographic features of AuTuMN that can be used to simulate births, ageing and deaths were not implemented, because the issues considered pertain to the short- to medium-term and the immediate implementation of control strategies, for which population demographics are less relevant.

##### 1.4 Clinical stratification

The age-stratified late exposed/incubation and both the early and late active disease compartments were further stratified into five "clinical" categories: *1)* asymptomatic, *2)* symptomatic ambulatory, never detected, *3)* symptomatic ambulatory, ever detected, *4)* ever hospitalised, never critical and *5)* ever critically unwell (Figure 2). The proportion of new infectious persons entering stratum 1 (asymptomatic) is age-dependent (as described in Table 4). The proportion of symptomatic patients (strata 2 to 5) ever detected (strata 3 to 5) is set through a parameter that represents the time-varying proportion of all symptomatic patients who are ever detected (the case detection rate, see Section 2). Of those ever symptomatic (strata 2 to 5), a time-constant but age-specific proportion is considered to be hospitalised (entering strata 4 or 5). Of those hospitalised (entering strata 4 or 5), a fixed proportion was considered to be critically unwell (entering stratum 5, Figure 3).

##### 1.5 Hospitalisation

For COVID-19 patients who are admitted to hospital, the sojourn time in the early and late active compartments is modified, superseding the default values of the sojourn times for these compartments, as indicated in Table 3. The point of admission to hospital is considered to be the transition from early to late active disease, such that the sojourn time in the late disease represents the period of time admitted to hospital. For patients admitted to ICU, admission to ICU occurs at this same transition point. For this group, the period of time hospitalised prior to ICU admission is estimated as a proportion of the early active period, such that the early active period represents both the period ambulatory in the community and the period in hospital prior to ICU admission.

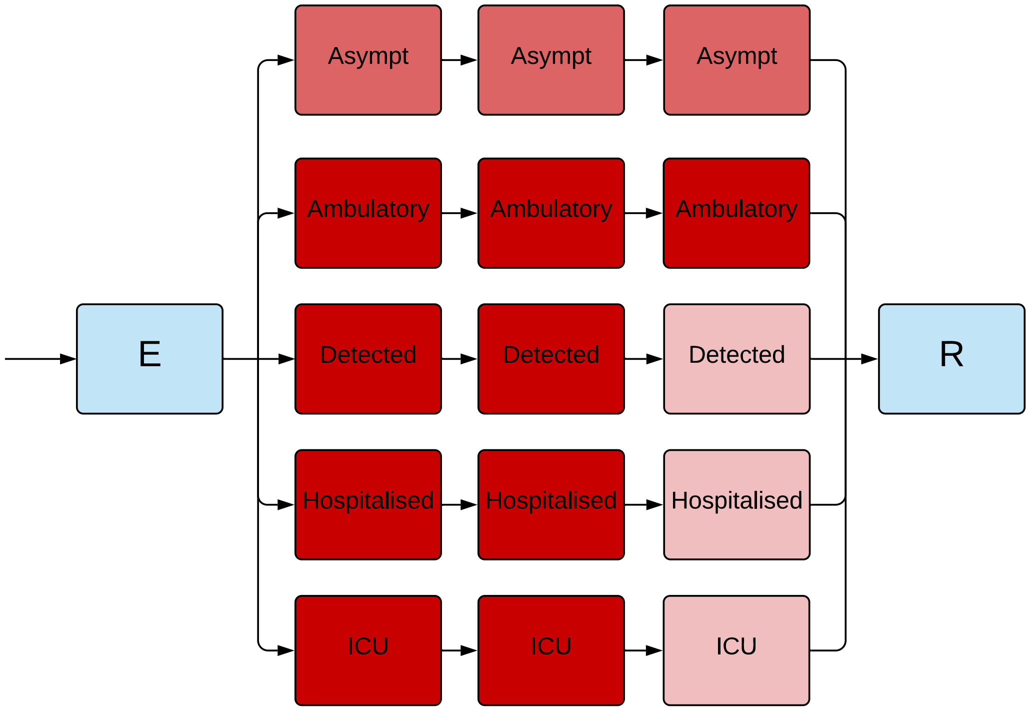

**Figure 2 – Illustration of the implementation of the clinical stratification.** Depth of pink/red shading indicates the infectiousness of the compartment. Typical parameter values presented, although the infectiousness of asymptomatic persons is varied in calibration.

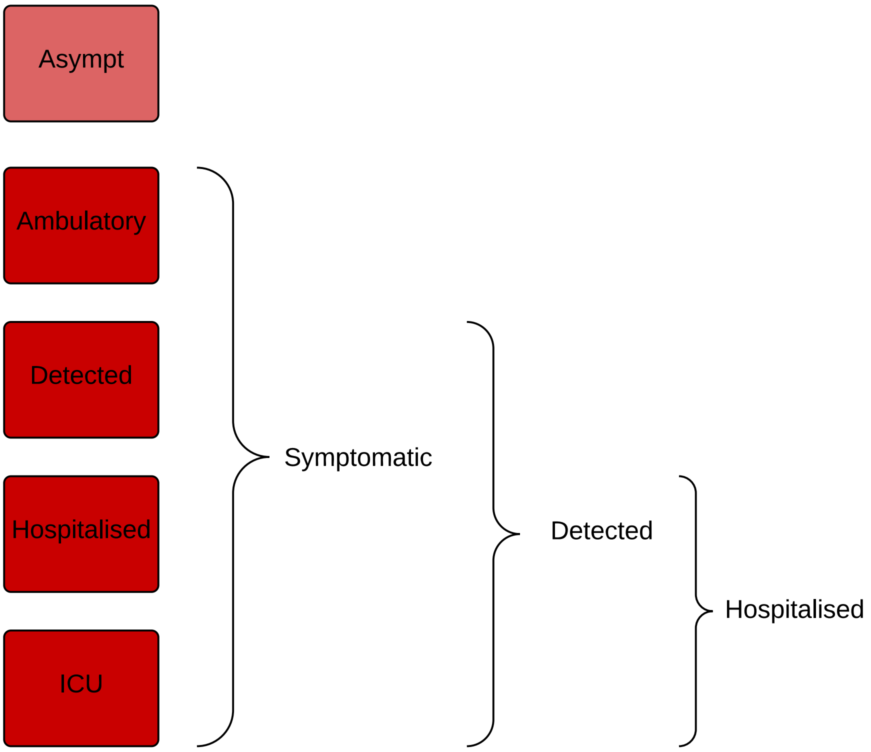

**Figure 3 – Illustration of the rationale for the clinical stratification.**

##### 1.6 Infectiousness

Asymptomatic persons are assumed to be less infectious per unit time active than symptomatic persons not undergoing case isolation (typically by around 50%, although this is varied in calibration/uncertainty analysis). Infectiousness is also decreased for persons who have been detected to reflect case isolation, and for those admitted to hospital or ICU to reflect infection control procedures (by 80% for both groups). Presymptomatic individuals are presumed to have equivalent infectiousness to those with early active COVID-19.

##### 1.7 Application of COVID-19-related death

Age-specific infection fatality rates (IFRs) were applied and distributed across strata 4 and 5, with no deaths typically applied to the first three strata. A ceiling of 50% is set on the proportion of those admitted to ICU (entering stratum 5) who die. If the infection fatality rate is greater than this ceiling, the proportion of critically unwell persons dying was set to 50%, with the remainder of the infection fatality rate then applied to the hospitalised proportion. Otherwise, if the infection fatality rate is less than half of the absolute proportion of persons critically unwell, the infection fatality rate is applied entirely through stratum 5 (such that the proportion of critically unwell persons dying in that age group becomes $<$50% and the proportion of stratum 4 dying is set to zero). In the event that the infection fatality rate for an age group is greater than the total proportion hospitalised (which is unusual, but could occur for the oldest age group under certain parameter configurations), the remaining deaths are assigned to the asymptomatic stratum. This approach was adopted for computational ease and is valid because the duration active for persons entering this stratum is the same as for the other non-hospitalised strata, such that the dynamics are identical to assigning the deaths to any of the first three strata. We used the age-specific IFRs previously estimated from age-specific death data from 45 countries and results from national-level seroprevalence surveys [4] as indicated in Table 4. We allowed IFRs to vary around the previously published point estimates in order to incorporate uncertainty and to allow the IFRs to differ from the settings in which they were estimated (see Calibration section).

| Clinical stratum | Stratum name | Pre-symptomatic | Early | Late |
|---|---|---|---|---|
| 1 | Asymptomatic | 0.5 | 0.5 | 0.5 |
| 2 | Symptomatic ambulatory never detected | 1 | 1 | 1 |
| 3 | Symptomatic ambulatory ever detected | 1 | 1 | 0.2 |
| 4 | Hospitalised never critical | 1 | 1 | 0.2 |
| 5 | Ever critically unwell | 1 | 1 | 0.2 |

**Table 1 – Illustration of the relative infectiousness of disease compartments by clinical stratification and stage of infection.** Typical parameter values displayed.

##### 1.8 Modelling Variants of Concern (VoC)

To consider the effects of VoC on infection dynamics and the vaccination programs, we explicitly simulated two competing strains to represent 1) the wild-type or ancestral virus, and 2) all VoC strains, where the VoC were assumed to be associated with increased transmissibility only. Therefore, we do not differentiate between different variants and assume the single VoC strain that is modelled represents all currently circulating strains. Susceptible individuals can be infected with either the wild-type or VoC strain and infectious individuals contribute to the force of infection with their respective infecting strain only. VoC strains are seeded into the model such that one additional person per day is infected with the VoC strain for a duration of ten days, with the time that this ten-day period commences varied during model calibration.

##### 1.9 Modelling vaccination

We stratified all model compartments as either vaccinated or unvaccinated and commenced simulations with an fully unvaccinated population. With vaccination roll-out, individuals in the susceptible and recovered compartments move from “unvaccinated” to “vaccinated” at a constant rate representing vaccine administration over time. The daily rate of vaccination is calculated from the vaccine coverage achieved over a given period of time is calculated as $rate_{vac} = \frac{-log(1-coverage)}{T}$, where coverage represents the targeted proportion of people vaccinated by the end of the roll-out period and $T$ represents the roll-out period.

##### 1.10 Modelling vaccine effects

Vaccination is assumed to have two mechanisms of effect: 1) prevention of infection and 2) protection against progressing to severe infection among those infected. A particular vaccine roll-out programme can be simulated to act through these two mechanisms simultaneously. The proportion of the effect that is attributed to preventing infection, $V_p = V_i V_e$ , where $V_p \in [0,1]$ is varied through model calibration, $V_i \in [0,1]$ is the infection prevention efficacy and $V_e \in [0,1]$ is the overall efficacy (that would be observed in clinical trials). If severity prevention efficacy is denoted $V_s$, since $V_e = V_i + V_s(1-V_i)$ it follows that $V_s = \frac{V_e(1-V_p)}{1-V_pVe}$ . For the component of the vaccine effect attributed to infection prevention, the infection risk of vaccinated individuals is reduced by $(1-V_i)$. Severity-preventing vaccination reduces the infection fatality rate (IFR) and the probability that an infected individual experiences symptomatic disease. Thus, the vaccine efficacy parameter pertaining to disease severity prevention modifies the splitting proportions of infected individuals between the different clinical categories and the rate of COVID-19-related mortality.

### 2 Case detection

##### 2.1 General approach

We calculate a time-varying case detection rate, being the proportion of all symptomatic cases (clinical strata 2 to 5) that are detected (clinical strata 3 to 5). This proportion is informed by the number of tests performed using the following formula:

$$CDR(time) = 1 - e^{-shape \times tests(time)}$$

*time* is the time in days from the 31st December 2019 and *tests(time)* is the number of tests per capita done on that date. To determine the value of the shape parameter, we solve this equation based on the assumption that a certain daily testing rate *tests(t)* is associated with a certain *CDR(t)*. Solving for *shape* yields:

$$shape = \frac{-log(1-CDR(t))}{tests(t)}$$

That is, if it is assumed that a certain daily per capita testing rate is associated with a certain proportion of symptomatic cases detected, we can determine *shape*. As this relationship is not well understood and unlikely to be consistent across all settings, we vary the *CDR* that is associated with a certain per capita testing rate during uncertainty/calibration. Given that the *CDR* value can be varied widely, the purpose of this is to incorporate changes in the case detection rate that reflect the empirical historical profile of changes in testing capacity over time.

### 3 Implementation of non-pharmaceutical interventions

A major part of the rationale for the development of this model was to capture the past impact of non-pharmaceutical interventions (NPIs) and produce future scenarios projections with the implementation or release of such interventions.

#### 3.1 Isolation and quarantine

For persons who are identified with symptomatic disease and enter clinical stratum 3, self-isolation is assumed to occur and their infectiousness is modified as described above. The proportion of ambulatory symptomatic persons effectively identified through the public health response by any means is determined by the case detection rate as described above.

#### 3.2 Community quarantine or “lockdown” measures

For all NPIs relating to reduction of human mobility or “lockdown” (i.e. all NPIs other than isolation and quarantine), these interventions are implemented through dynamic adjustments to the age-assortative mixing matrix. The baseline mixing matrices of Prem et al. [3] are synthetic and do not represent direct observations or reports from surveys (in the case of the 144 countries to which they were extrapolated from observations in the eight “POLYMOD” countries of Western Europe). Although synthetic, the matrices are contextualised to national demographic information, including country-specific data that include household size, workforce participation and school enrolment. Further, the matrices presented are easily machine-readable and appear to be plausible representations of contact structures within these countries.

The matrices also have the major advantage of allowing for disaggregation of total contact rates by location, i.e. home, work, school and other locations. This disaggregation allows for the simulation of various NPIs in the local context by dynamically varying the contribution of each location to reflect the historical implementation of the interventions.

The corresponding mixing matrix (denoted $C_0$) is presented using the standard convention that a row represents the average number of age-specific contacts per day for a contact recipient of a given age-group. In other words, the element $C_{0i,j}$ is the average number of contacts per day that an individual of age-group $_j$ makes with individuals of age-group $_i$.

This matrix results from the summation of the four location-specific contact matrices provided by Prem et al.: $C_0 = C_H + C_S + C_W + C_L$, where $C_H$, $C_S$, $C_W$ and $C_L$ are the age-specific contact matrices associated with households, schools, workplaces and other locations, respectively.

In our model, the contributions of the matrices $C_S$, $C_W$ and $C_L$ vary with time such that the input contact matrix can be written:

$$C(t) = C_H + s(t)^2 C_S + w(t)^2 C_W + l(t)^2 C_L$$

The modifying functions are each squared to capture the effect of the mobility changes on both the infector and the infectee in any given interaction that could potentially result in transmission. The modifying functions incorporate both macro-distancing and microdistancing effects, depending on the location.

##### 3.3 School closures/re-openings

Reduced attendance at schools is represented through the function $s(t)$, which represents the proportion of all school students currently attending on-site teaching. If schools are fully closed, $s(t) = 0$ and $C_S$ does not contribute to the overall mixing matrix $C(t)$. $s(t)$ is calculated through a series of estimates of the proportion of students attending schools, to which a smoothed step function is fitted. Note that the dramatic changes in this contribution to the mixing matrix with school closures/re-openings is a more marked change than is seen with the simulation of policy changes in workplaces and other locations (which are determined by empiric data and so do not vary so abruptly and do not fall to zero).

##### 3.4 Workplace closures

Workplace closures are represented by quadratically reducing the contribution of workplace contacts to the total mixing matrix over time. This is achieved through the scaling term $w(t)^2$ which modifies the contribution of $C_W$ to the overall mixing matrix $C(t)$. The profile of the function $w(t)$ is set by fitting a polynomial spline function to Google mobility data for workplace attendance (Table 2).

##### 3.5 Community-wide movement restriction

Community-wide movement restriction (or "lockdown") measures are represented by proportionally reducing the contribution of the other locations contacts to the total mixing matrix over time. This is achieved through the scaling term $l(t)^2$ which modifies the contribution of $C_L$ to the overall mixing matrix $C(t)$. The profile of the function $l(t)$ is set by fitting a polynomial spline function to an average of Google mobility data for various locations, as indicated in Table 2.

##### 3.6 Household contacts

The contribution of household contacts to the overall mixing matrix $C(t)$ is fixed over time. Although Google provides mobility estimates for residential contacts, the nature of these data are different from those for each of the other Google mobility types in that they represent the time spent in that location rather than the duration. The daily frequency with which people attend their residence is likely to be close to one and we considered that household members likely have a daily opportunity for infection with each other household member. Therefore, we did not implement a function to scale the contribution of household contacts to the mixing matrix with time.

##### 3.7 Microdistancing

Interventions other than those that prevent people coming into contact with one another are thought to be important to COVID-19 transmission and epidemiology, such as maintaining interpersonal physical distance and the wearing of face coverings. We therefore implemented a "microdistancing" function to represent reductions in the rate of effective contact that is not attributable to persons visiting specific locations and

| Prem "location" | Approach | Google mobility types |
|---|---|---|
| School | Policy response | Not applicable |
| Household | Constant | Not applicable |
| Workplace | Google mobility | Workplace |
| Other locations | Google mobility | Unweighted average of:<br>• Retail and recreation<br>• Grocery and pharmacy<br>• Parks<br>• Transit stations |

**Table 2** – **Mapping of Google mobility data to contact locations** (as defined by Prem et al.)

so is not captured through Google mobility data. This microdistancing function reduces the values of all elements of the mixing matrices by a certain proportion. These time-varying functions multiplicatively scale the location-specific contact rate modifiers $s(t)$, $w(t)$ and $l(t)$.

### 4 Parameters

#### 4.1 Non-age-stratified parameters

| Parameter | Value | Rationale |
|---|---|---|
| Incubation period | Calibration parameter, truncated normal distribution, mean 5.5 days | Estimates of the incubation period have included 5.1 days, 5.2 days and 4.8 days [5] [6] [7] [8]. A systematic review [2] found that data are best fitted by a log-normal distribution (mean 5.8 days, CI 5.0 to 6.7, median 5.1 days). Our systematic review [9] found that estimates of the mean incubation period have varied from 3.6 to 7.4 days. |

Continuation of Table 3

| Parameter | Value | Rationale |
|---|---|---|
| Proportion of incubation period infectious | 50% | Infectiousness is considered to be present throughout a considerable proportion of the incubation period, based on analyses of confirmed source-secondary pairs [10] and early findings that the incubation period was similar to the serial interval [5]. The study of source-secondary pairs was also the primary reference cited by a review of the infectious period that identified studies that quantified the pre-symptomatic period, which concluded that the median pre-symptomatic period could range from less than one to four days [11]. |
| Active period (regardless of detection/isolation, for clinical strata 1 to 3) | Calibration parameter, truncated normal distribution, mean 8 days | This quantity is difficult to estimate, given that identified cases are typically quarantined. Studies in settings of high case ascertainment and an effective public health response have suggested a duration of greater than 5.5 days [8]. PCR positivity, which may continue for up to two to three weeks from the point of symptom onset [10] [11], is difficult to interpret and does not necessarily indicate infectiousness. Consistent with these findings, the duration infectious for asymptomatic persons has been estimated at 6.5 to 9.5 days [11] (although in our model, this would include the pre-symptomatic infectious period). |
| Proportion of infectious period before isolation or hospitalisation can occur | 0.333 | Assumed |
| Disease duration prior to admission for hospitalised patients not critically unwell (i.e. early active sojourn time, stratum 4) | 7.7 days | Mean value from ISARIC cohort, as reported on 4$^{th}$ October 2020 in Table 6 [12], and similar to the expected mean from earlier reports from ISARIC [13]. This cohort represents high-income countries better than low and middle-income countries, with the United Kingdom contributing data on the greatest number of patients, followed by France. Earlier estimates of this quantity from China included 4.4 days [5]. |
| Duration of hospitalisation if not critically unwell (late active sojourn time, stratum 4) | 12.8 days | Mean value from the ISARIC cohort, as reported on 4$^{th}$ October 2020 in Table 6 [12]. |

Continuation of Table 3

| **Parameter** | **Value** | **Rationale** |
|---|---|---|
| ICU duration (late active sojourn time, stratum 5) | 10.5 days | Mean duration of stay in ICU/HDU from ISARIC cohort for patients with complete data, as reported on 10$^{th}$ October 2020 Table 6 [12]. Many other studies reporting on the average duration of ICU stay suffer from right-truncation issues, often estimating 7-10 days length of stay. |
| Duration of time prior to ICU for patients admitted to ICU | 10.5 days | Calculated as the sum of the time from symptom onset to hospital admission (7.7 days above) plus the duration from hospital admission to ICU admission reported by October ISARIC report (2.8 days) [12]. |
| Relative infectiousness of asymptomatic persons (per unit time with active disease) | 0.5 | Assumed |
| Relative infectiousness of persons admitted to hospital or ICU | 0.2 | Assumed |
| Relative infectiousness of identified persons in isolation | 0.2 | Assumed |
| Proportion of hospitalised patients ever admitted to ICU | 0.17 | Assumed |

**Table 3** – **Universal (non-age-stratified) model parameters.** Point estimates are used as model parameters except where ranges are indicated in calibration parameter table below in calibration table.

##### *4.2 Age-specific parameters*

| **Age group (years)** | **Clinical fraction[a]** | **Relative susceptibility to infection** | **Infection fatality rate** | **Proportion of symptomatic patients hospitalised** |
|---|---|---|---|---|
| 0 to 4 | 0.29 | 0.36 | $3 \times 10^{-5}$ | 0.0777 |
| 5 to 9 | 0.29 | 0.36 | $1 \times 10^{-5}$ | 0.0069 |
| 10 to 14 | 0.21 | 0.36 | $1 \times 10^{-5}$ | 0.0034 |
| 15 to 19 | 0.21 | 1 | $3 \times 10^{-5}$ | 0.0051 |
| 20 to 24 | 0.27 | 1 | $6 \times 10^{-5}$ | 0.0068 |
| 25 to 29 | 0.27 | 1 | $1.3 \times 10^{-4}$ | 0.0080 |
| 30 to 34 | 0.33 | 1 | $2.4 \times 10^{-4}$ | 0.0124 |
| 35 to 39 | 0.33 | 1 | $4.0 \times 10^{-4}$ | 0.0129 |
| 40 to 44 | 0.40 | 1 | $7.5 \times 10^{-4}$ | 0.0190 |
| 45 to 49 | 0.40 | 1 | $1.21 \times 10^{-3}$ | 0.0331 |
| 50 to 54 | 0.49 | 1 | $2.07 \times 10^{-3}$ | 0.0383 |
| 55 to 59 | 0.49 | 1 | $3.23 \times 10^{-3}$ | 0.0579 |
| 60 to 64 | 0.63 | 1 | $4.56 \times 10^{-3}$ | 0.0617 |
| 65 to 69 | 0.63 | 1.41 | $1.075 \times 10^{-2}$ | 0.1030 |
| 70 to 74 | 0.69 | 1.41 | $1.674 \times 10^{-2}$ | 1.072 |
| 75 and above | 0.69 | 1.41 | $5.748 \times 10^{-2}$, [b] | 0.0703 |
| Source/ rationale | Model fitting to age-distribution of early cases in China, Italy, Japan, Singapore, South Korea and Canada taken from upper-left panel of Figure 2b of [14]. | Conversion of odds ratios presented in Table S15 of Zhang et al. 2020 to relative risks using data presented in Table S14 of the same study [15].[c] | Estimated from pooled analysis of data from 45 countries from Table S3 of O'Driscoll et al [4]. Values consistent with previous estimates using serosurveys performed in Spain [16]. | Estimates from the Netherlands as the first wave of infections declined from 4th May to 21st July [17]. |

**Table 4** – **Age-stratified parameter values.** Age-stratified parameters not varied during calibration, or varied through a common adjuster parameter.
[a] Proportion of incident cases developing symptoms.
[b] Weighted average of IFR estimates for 70 to 79 and 80 and above age groups.
[c] Note the relative magnitude of these values are similar to those estimated by the analysis we use to estimate the age-specific clinical fraction.[14]

### 5 Calculation of outputs

#### 5.1 Incidence

Incidence is calculated as any transitions into the early active compartment (*"I"*).

#### 5.2 Hospital occupancy

This is calculated as the sum of three quantities:

1. All persons in the late active compartment in clinical stratum 4, representing those admitted to hospital but never critically unwell.
2. All persons in the late active compartment in clinical stratum 5, representing those currently admitted to ICU.
3. A proportion of the early active compartment in clinical stratum 5, representing those who will be admitted to ICU at a time in the future. This proportion is calculated as the quotient of *1)* the difference between the pre-ICU period and the pre-hospital period for clinical stratum 4, divided by *2)* the total pre-ICU period. That is, a proportion of the pre-ICU period is considered to represent patients in hospital who have not yet been admitted to ICU.

#### 5.3 ICU occupancy

This is calculated as all persons in the late active compartment in clinical stratum 5.

#### 5.4 Seropositive proportion

This is calculated as the proportion of the population in the recovered (*"R"*) compartment. Although very similar numerically to the attack rate, persons who died of COVID-19 are not included in the denominator.

#### 5.5 COVID-19-related mortality

This is calculated as all transitions representing death, exiting the model. This is implemented as depletion of the late active compartment.

##### 5.6 Notifications

Local case notifications are calculated as transitions from the early to the late active compartment for clinical strata 3 to 5.

#### 6 Calibration

##### 6.1 General approach

The model was calibrated using an adaptive Metropolis (AM) algorithm. In particular, we used the algorithm based on adaptive Gaussian proposal functions proposed by Haario *et al.* to sample parameters from their posterior distributions [18]. We ran seven independent AM chains and combined the samples of the seven chains to project epidemic trajectories over time. The definitions of the prior distributions and the likelihood are detailed as follows.

##### 6.2 Likelihood function

Likelihood functions are derived from comparing model outputs to target data at each time point nominated for calibration. The dispersion parameters of these distribution are included as calibration parameters and varied during the calibration approach to improve calibration efficiency.

##### 6.3 Variation of symptomatic proportions and proportion hospitalised

Whether age-specific proportions of symptomatic individuals are significantly different in low- and middle-income settings from those in high-income settings remains highly uncertain. For this application to the Malaysia, we incorporated uncertainty around the age-specific proportions of symptomatic individuals by applying an uncertainty adjuster:

$$s_i^* = \frac{s_i \times \gamma}{s_i(\gamma - 1) + 1},$$

where $s_i^*$ is the modelled symptomatic proportion for age group $i$, $s_i$ is the point estimate reported by Davies *et al.* for the IFR of age group $i$ [14], and $\gamma$ is the associated uncertainty adjuster varied during model calibration.

Similarly, we incorporated uncertainty around the age-specific proportions hospitalised by applying an uncertainty adjuster:

$$h_i^* = \frac{h_i \times \beta}{h_i(\beta - 1) + 1},$$

where $h_i^*$ is the modelled hospitalised proportion for age group $i$, $h_i$ is the point estimate, and $\beta$ is the associated uncertainty adjuster varied during model calibration.

##### 6.4 Calibration parameters

| **Parameter name** | **Distribution type** | **Distribution parameters** |
|---|---|---|
| Incubation period | Truncated normal | Mean 5.5 days, standard deviation 0.97 days, truncation $<$1 day |
| Infectious period (for clinical strata 1 to 3) | Truncated normal | Mean 6.5 days, standard deviation 0.77 days, truncation $<$4 days |
| Risk of infection per contact | Uniform | 0.015 to 0.06 |
| Proportion of symptomatic cases that would be detected with a daily per capita testing rate of one test per ten thousand population | Uniform | Range 0.02 to 0.1 |
| Infectious seed | Uniform | Range 30 to 200 |

Continuation of Table 5

| Parameter name | Distribution type | Distribution parameters |
|---|---|---|
| Adjuster applied to age-specific proportion of infections leading to symptoms (“Clinical fraction”) | Uniform | 0.2 to 2 |
| Adjuster applied to age-specific proportion of symptomatic infections leading to hospitalisations | Uniform | 0.8 to 2 |
| Relative infectiousness of asymptomatic persons (per unit time with active disease) | Uniform | 0.15 to 0.4 |
| Increased transmissibility of VoC strains | Uniform | 1.2 to 2.0 |
| Start time of VoC emergence | Uniform | 300 to 400 |

**Table 5 – Epidemiological calibration parameters.**

##### *6.5 Calibration targets*

We calibrated the Malaysia national model to three targets; case notifications, intensive care unit (ICU) occupancy and infection-related deaths. Although the ICU occupancy and mortality data was too sparse for the regional models, aggregating the data across space and to weekly national values allowed us to include these two targets (ICU occupancy and mortality) in the national model. Therefore the regional models are calibrated to only the notification data.

tabularx

#### 7 Ordinary differential equations

For the clearest description of the model, we refer the reader to our code repository, because our object-oriented approach to software development is intended to be highly transparent and readable. For those who prefer dynamical systems such as those presented in the form of ordinary differential equations, we present the following.

$$\frac{dS_a}{dt} = -\lambda_a(t) \times \sigma_a \times S_a$$
$$\frac{dE_a}{dt} = \lambda_a(t) \times \sigma_a \times S_a - \alpha E_a$$
$$\frac{dP_{a,c}}{dt} = p_{a,c}(t) \times \alpha E_a - \nu P_{a,c}$$
$$\frac{dI_{a,c}}{dt} = \nu P_{a,c} - \gamma_c I_{a,c}$$
$$\frac{dL_{a,c}}{dt} = \gamma_c I_{a,c} - \delta_{a,c} L_{a,c} - \mu_{a,c} L_{a,c}$$
$$\frac{dR_a}{dt} = \sum_c \delta_{a,c} L_{a,c}$$

where

$$\lambda_a = \beta \left[ \sum_{j,c} \frac{\varepsilon \times P_j}{N_j} \times C_{a,j}(t) + \sum_{j,c} \frac{I_{j,c} \times \iota_c + L_{j,c} \times \kappa_c}{N_j} \times C_{a,j}(t) \right]$$
$$\sum_c p_{a,c}(t) = 1, \forall t \in \mathbb{R}$$
$$\mathbf{C}_0 = \mathbf{C}_H + \mathbf{C}_S + \mathbf{C}_W + \mathbf{C}_L$$
$$\mathbf{C}(t) = h(t) \times \mathbf{C}_H + s(t) \times \mathbf{C}_S + w(t) \times \mathbf{C}_W + l(t) \times \mathbf{C}_L$$
$$l(t) = \frac{re(t) + gr(t) + pa(t) + tr(t)}{4}$$

| Symbol | Explanation |
|---|---|
| $S$ | Persons susceptible to infection |
| $E$ | Persons in the non-infectious incubation period |
| $P$ | Persons in the incubation period |
| $I$ | Persons in the early active disease period, before isolation or hospitalisation may occur |
| $L$ | Persons in the late active disease period, after isolation or hospitalisation may have occurred |
| $R$ | Persons in the recovered period, from which re-infection cannot occur |

| Symbol | Explanation |
|---|---|
| $t$ | Time |
| $_a$ | Compartment of age group a |
| $_c$ | Compartment of clinical stratification c |
| $\sigma$ | Relative susceptibility to infection |
| $\alpha$ | Rate of progression from non-infectious to infectious incubation period |
| $\nu$ | Rate of progression from infectious incubation to early active disease |
| $\gamma$ | Rate of progression from early active disease to late active disease |
| $\mu$ | Rate of disease-related death |
| $\varepsilon$ | Relative infectiousness of pre-symptomatic compartment |
| $\iota$ | Clinical stratification infectiousness vector for early active compartment |
| $\kappa$ | Clinical stratification infectiousness vector for late active compartments |
| $\beta$ | Probability of infection per contact between an infectious and susceptible individual |
| $j$ | Infectious populations |
| $p$ | Proportion progressing to each clinical stratification |

| Symbol | Explanation |
|---|---|
| $\mathbf{C}$ | Mixing matrix |
| $\mathbf{H}$ | Household contribution to mixing matrix |
| $\mathbf{W}$ | Workplace contribution to mixing matrix |
| $\mathbf{O}$ | Other locations contribution to mixing matrix |
| $\mathbf{S}$ | Schools contribution to mixing matrix |
| $l$ | Other locations macrodistancing function of time |
| $w$ | Function fit to Google mobility data for workplaces |
| $s$ | Function fit to Google mobility data for schools |
| $re$ | Function fit to Google mobility data for retail and recreation |
| $gr$ | Function fit to Google mobility data for grocery and pharmacy |
| $pa$ | Function fit to Google mobility data for parks |
| $tr$ | Function fit to Google mobility data for transit stations |

### 8 Supplemental figures to main text

#### 8.1 Supplemental Figures

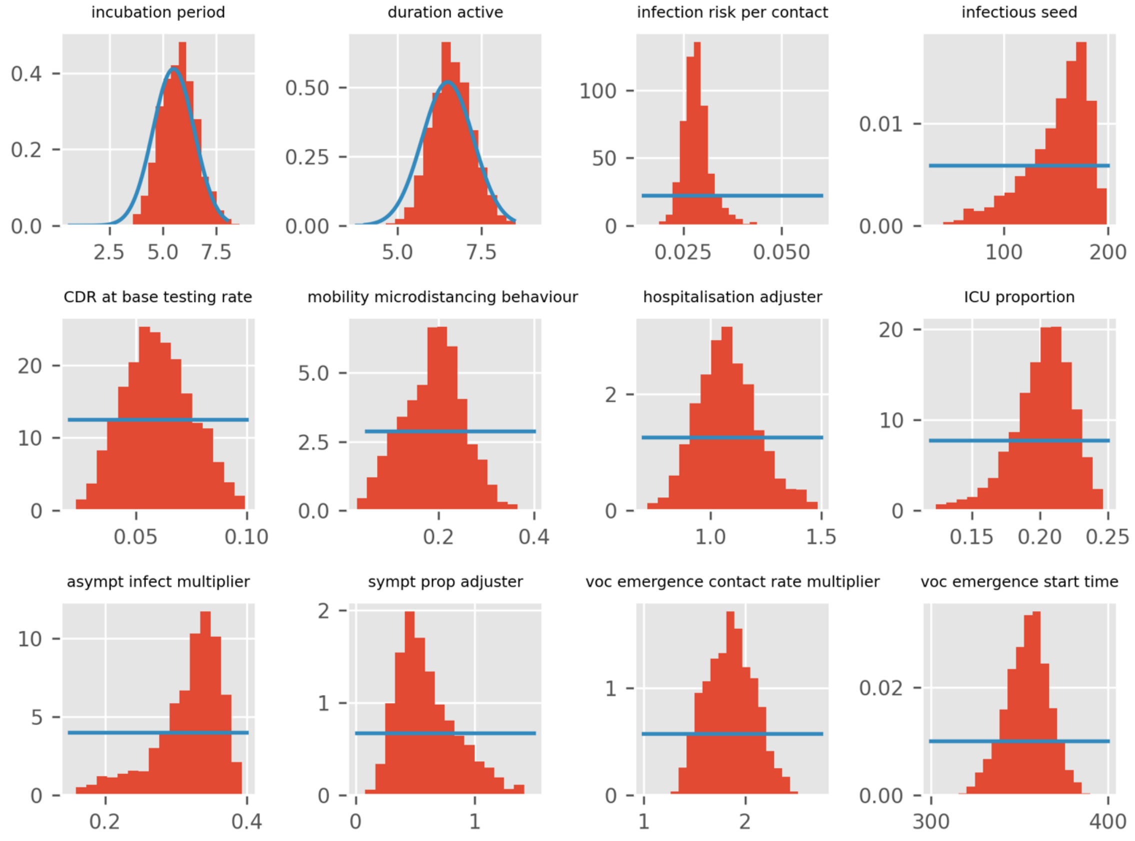

**Figure 4 – Histograms of prior (blue lines) and posterior (red bars) distributions for epidemiological parameters for Malaysia. In the VoC emergence start time the lower bound of the prior (300 days) corresponds to Nov 27, 2020 and the upper bound (400 days) corresponds to Feb 03, 2021.**

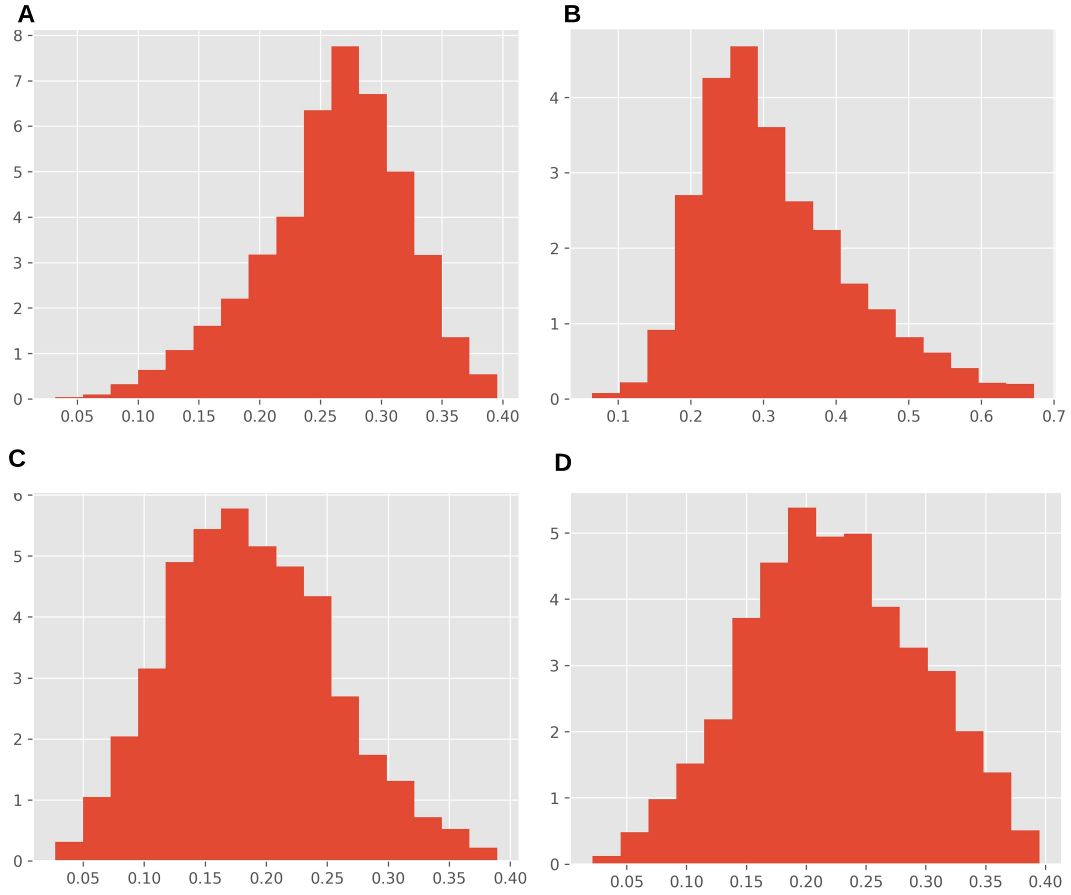

**Figure 5 – Posterior distributions of the micro-distancing parameter for the four regional models, Kuala Lumpur (A), Johor (B), Penang (C) and Selangor (D).**

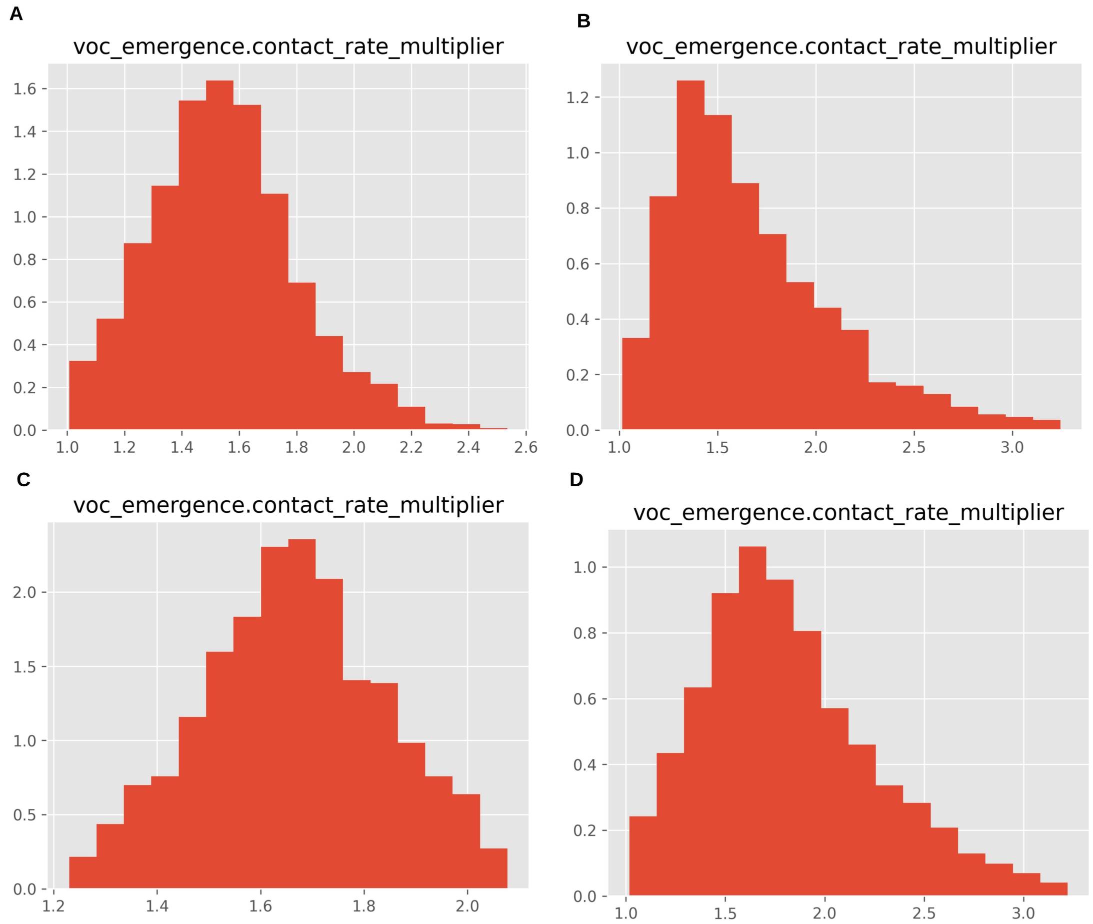

**Figure 6 – Posterior distributions of the increased transmissibility of VoC strains for the four regional models, Kuala Lumpur (A), Johor (B), Penang (C) and Selangor (D).**

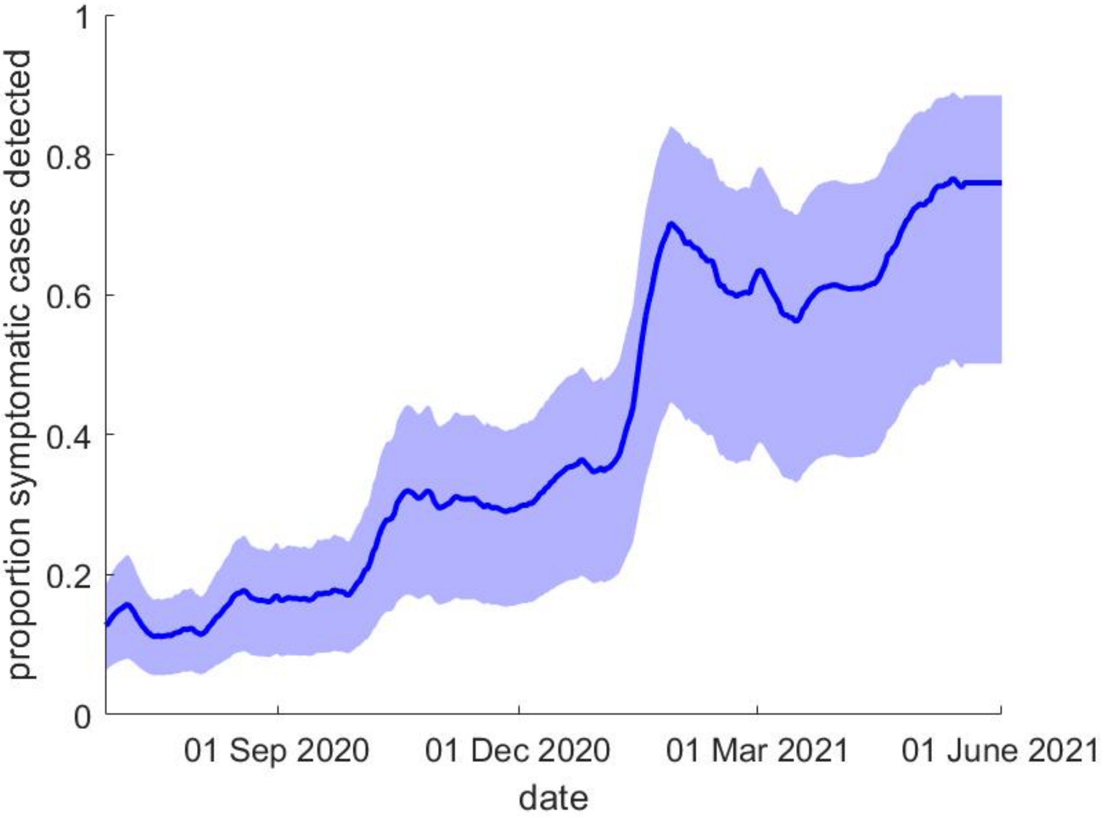

**Figure 7 – Model estimated proportion of symptomatic cases for the Malaysia nationwide model.**

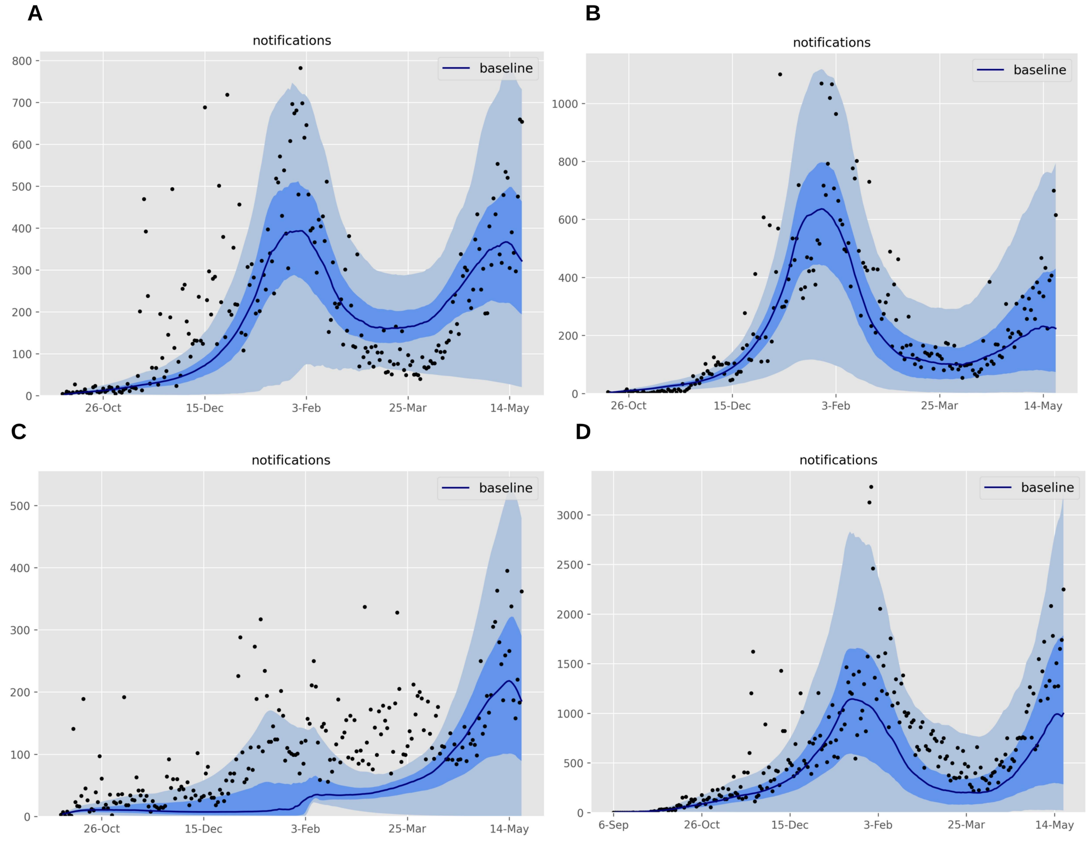

**Figure 8 – Model calibration for the four regional models (A) Kuala Lumpur; (B) Johor; (C) Penang and; (D) Selangor. The solid line is the median and the dark shading represents 25th to 75th centile credible interval; light shading 2.5th to 97.5th centile credible interval.**

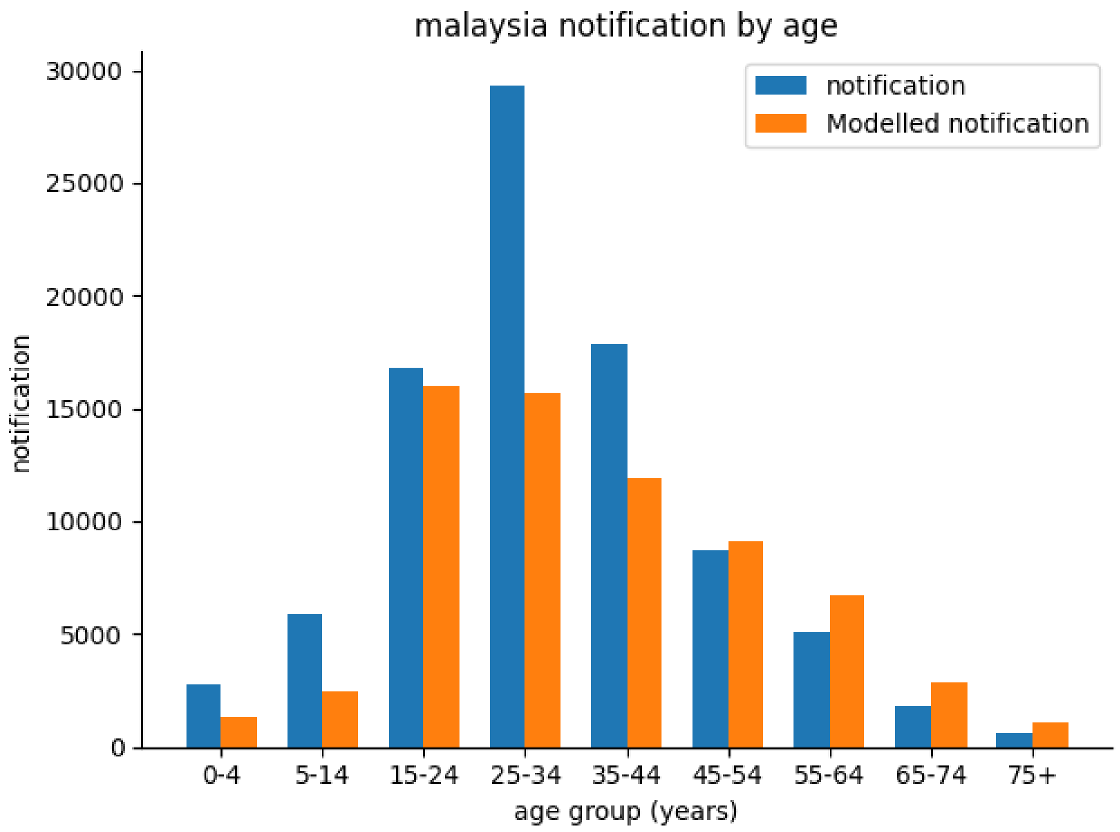

**Figure 9 – Modelled (orange bars) versus reported (blue bars) cumulative cases by age group for the Malaysia national model.**

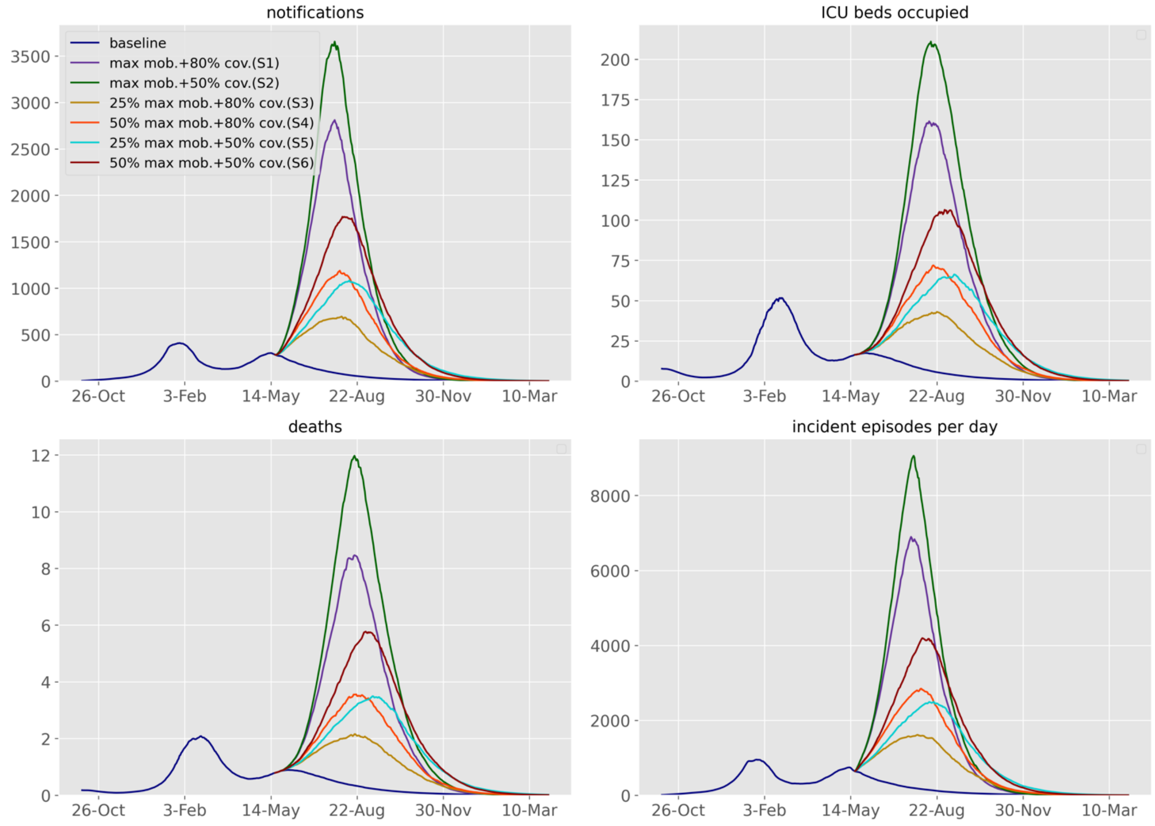

**Figure 10 – Future projections of the COVID-19 epidemic in Kuala Lumpur under various response scenarios and baseline. Upper-left, daily number of notifications; upper-right, ICU beds occupied; lower-left, daily number of COVID-19-related deaths; lower-right, incidence. For better visualisation, the median fits and projections are shown without uncertainty bounds. The time before the intervention start is shown for comparison with the previous epidemic**

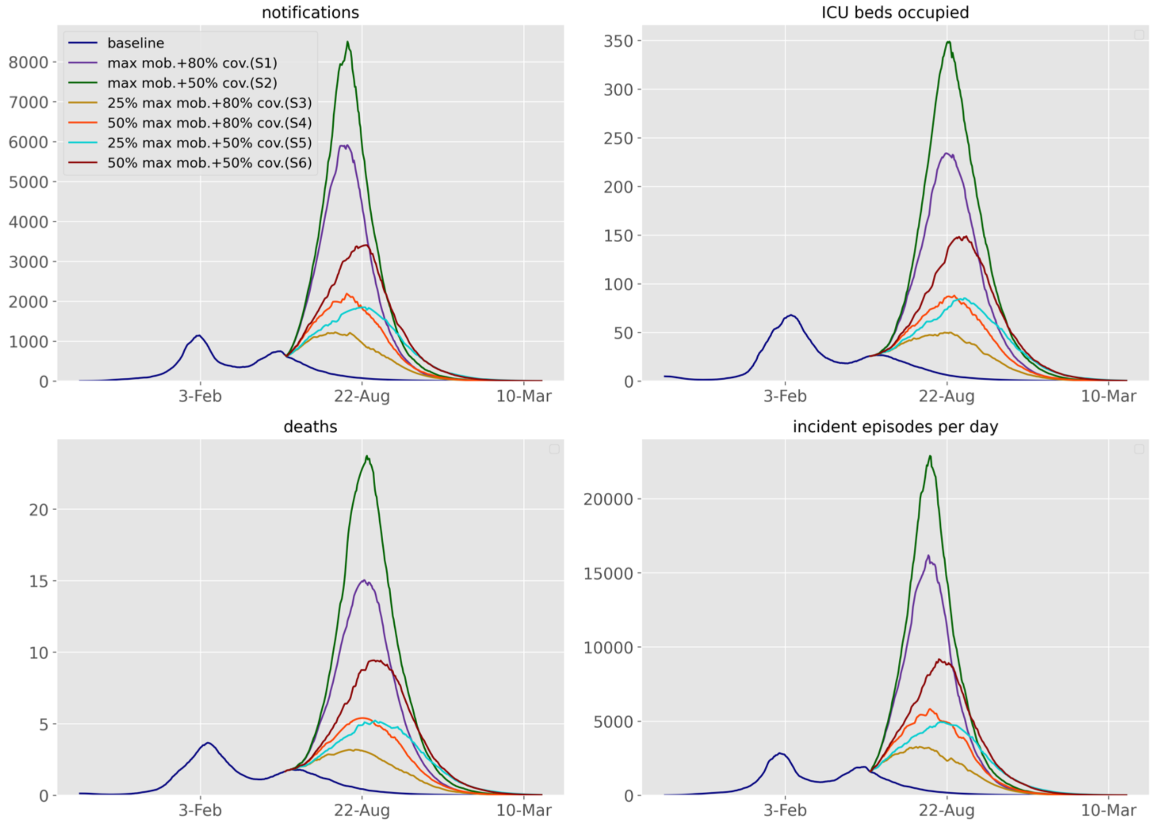

**Figure 11 – Future projections of the COVID-19 epidemic in Selangor under various response scenarios and baseline. Upper-left, daily number of notifications; upper-right, ICU beds occupied; lower-left, daily number of COVID-19-related deaths; lower-right, incidence. For better visualisation, the median fits and projections are shown without uncertainty bounds. The time before the intervention start is shown for comparison with the previous epidemic.**

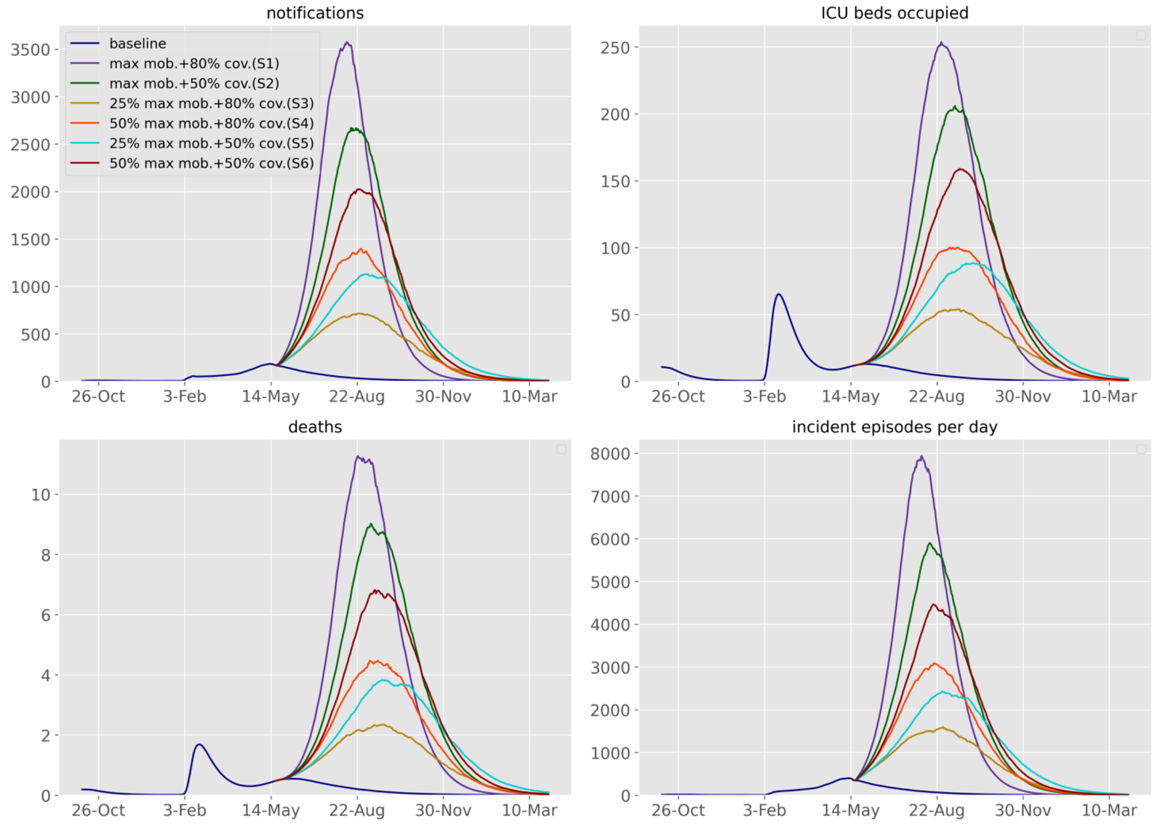

**Figure 12 – Future projections of the COVID-19 epidemic in Penang under various response scenarios and baseline. Upper-left, daily number of notifications; upper-right, ICU beds occupied; lower-left, daily number of COVID-19-related deaths; lower-right, incidence. For better visualisation, the median fits and projections are shown without uncertainty bounds. The time before the intervention start is shown for comparison with the previous epidemic.**

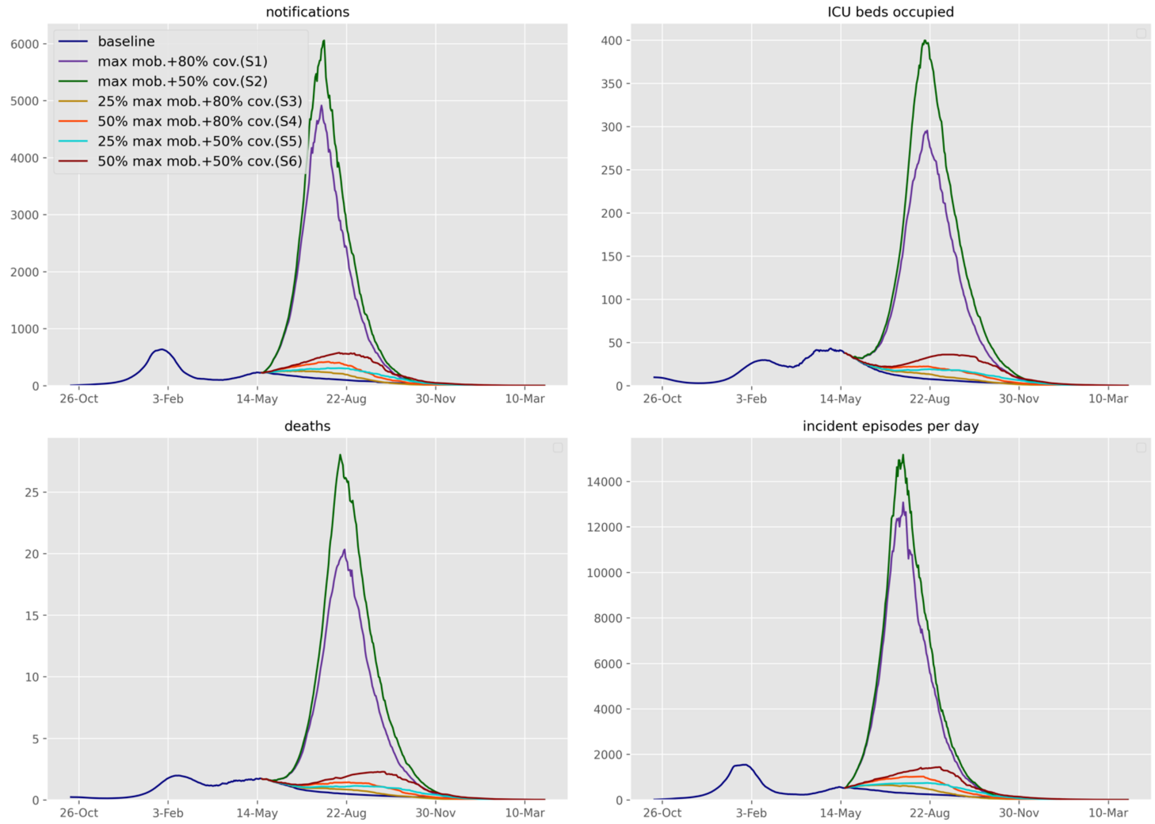

**Figure 13 – Future projections of the COVID-19 epidemic in Johor under various response scenarios and baseline. Upper-left, daily number of notifications; upper-right, ICU beds occupied; lower-left, daily number of COVID-19-related deaths; lower-right, incidence. For better visualisation, the median fits and projections are shown without uncertainty bounds. The time before the intervention start is shown for comparison with the previous epidemic.**
